# Multidimensional dietary patterns and their joint associations with intersecting sociodemographic characteristics among adults in Canada: a cross-sectional study

**DOI:** 10.1101/2024.12.11.24318868

**Authors:** Joy M. Hutchinson, Dylan Spicker, Benoît Lamarche, Michael Wallace, Mélina Côté, Abel Torres-Espín, Sharon I. Kirkpatrick

**Author notes:** **Sources of support:** This project was funded by an Ontario Ministry of Research and Innovation Early Researcher Award held by SIK. JMH was funded by a Vanier Canada Graduate Scholarship (Social Sciences and Humanities Research Council). **Corresponding author:** Joy Hutchinson PhD, RD.

## Abstract

**Background:** Dietary patterns consist of multiple interrelated components, while individuals have numerous characteristics that may jointly influence dietary patterns. Studies to assess associations between sociodemographic characteristics and dietary patterns typically do not consider this complexity.

**Objective:** The objective of this study was to examine joint relationships between dietary patterns and sociodemographic characteristics among adults in Canada.

**Methods:** 24-hour dietary recall data for adults ≥18 years were drawn from the 2015 Canadian Community Health Survey Nutrition (n=14 097). Three mixed graphical models were developed to explore networks of sociodemographic characteristics, dietary components, and sociodemographic characteristics and dietary components together. Networks included 30 log-transformed food groups (grams), sex, age, household food security status, income, employment status, education, geographic region, and smoking status. Results are expressed as (edge weight; [95% CI]).

**Results:** The strongest pairwise relationships were observed among dietary components and among sociodemographic characteristics. Positive linear relationships were observed among vegetable groupings; for example, between green and orange vegetables (0.12; [0,08, 0.16]). Negative relationships were observed among subgroups of each of animal foods, beverages, and grains; for example, between refined and whole grains (−0.30; [−0.33, −0.26]). In the model including dietary components and sociodemographic characteristics, age was associated with grains (other) (−0.12; [−0.16, −0.09]), coffee/tea (0.21; 95% CI [0.17, 0.24]), and whole grains (0.12; [0.08, 0.15]). Sex was associated with sweet beverages (0.11; [0.06, 0.17]), alcohol (0.18; [0.13, 0.24]), cured meat (0.20; [0.15, 0.26]), and red meat (0.16; [0.11, 0.21]).

**Conclusions:** In some cases, pairwise relationships between dietary components suggest displacement, for example, of whole grains by refined grains. Age and sex were the characteristics most strongly connected to dietary components.

**Statement of significance:** Exploring joint relationships between intersecting sociodemographic characteristics and multidimensional dietary patterns can assist with better understanding dietary heterogeneity to inform policies and programs that support healthy eating.

## Introduction

While nutrition and health research has historically been rooted in the examination of single dietary components, interest has shifted from researching single components to broader dietary patterns (1–3). Humans typically do not eat nutrients or foods in isolation, but in combination as part of the total diet (4). In general, the overall dietary pattern, with potential multidimensional synergistic or antagonistic interactions among dietary components, contributes more strongly to the development of diet-related chronic disease than single foods or nutrients (5–7). For example, evidence suggests that eating a variety of berries in combination may be more beneficial for the prevention of cancer than when consumed individually (8). More broadly, understanding the combinations in which foods are consumed as part of population dietary patterns could help to illuminate relationships with health and lead to nutrition policies and dietary guidance with the capacity to improve overall dietary patterns.

Much dietary patterns research has relied on *a priori* approaches, using expert knowledge and researcher hypotheses to create indices and scores indicative of dietary patterns associated with health (9–11). However, *a priori*-identified indices may not be fully representative of the range of potentially high quality dietary patterns among diverse populations (12–14). Further, using *a priori* approaches, multidimensional inputs are typically compressed to a unidimensional composite score, which is readily interpretable but does not capture complex interactions among different dietary components within the pattern. *A posteriori* approaches, including multidimensional analysis methods such as principal component analysis or factor analysis, have also provided opportunities to investigate dietary patterns and do not rely on researcher hypotheses (4,15). For instance, principal component analysis and factor analysis have been employed to characterize combinations of key components that constitute dietary patterns (15). These approaches are useful for gaining insights into the dietary patterns of different populations and population subgroups, but also typically provide condensed metrics (e.g., factor loadings) based on multidimensional inputs (4,16). To date, the methods predominantly used in the literature limit examinations of dietary synergy (2,17,18).

Not only are dietary patterns inherently complex in terms of the combinations of foods consumed, but they are shaped by sociodemographic characteristics and other factors such as culture (19–23). Differences in dietary intake have been observed in relation to several characteristics, including age, sex, income, education, household food security status, and smoking status (19,21–25). However, most research has not considered the potential combined effects of intersecting sociodemographic characteristics and their relationships with dietary patterns. Sociodemographic characteristics may intersect in ways that are compounded by forces of oppression to shape the dietary patterns that people are able to consume, as postulated by intersectionality theory (26–28). Research considering these intersections suggests that sociodemographic characteristics jointly contribute to dietary intake (29). For example, a study of adults in Canada found that when racial identity and perceived income adequacy were considered separately, only perceived income adequacy was meaningfully associated with diet quality (29). However, when the intersection of these characteristics was included, an association with diet quality was observed, suggesting the joint consideration of these characteristics is relevant (29). In the realm of precision nutrition, accounting for intersecting sociodemographic characteristics is important to inform strategies to improve dietary patterns (30,31). Considering intersections among characteristics can also have important implications for population-level dietary recommendations, such as the development of appropriate, tailored dietary guidance.

Undirected probabilistic graphical models offer the opportunity to examine how individual dietary components are interrelated with one another and with other factors (32,33). Undirected graphical models identify joint probability distributions in which the absence of a link between variables means that those variables are independent conditional on other variables, and a link represents associations that remain unexplained after accounting for all other variables (34,35). Probabilistic graphical models are well-suited to investigating multidimensionality because they can identify conditional relationships between the foods and food groupings that comprise high-dimensional dietary patterns (32,33,35–40). In other words, probabilistic graphical models allow for the examination of joint relationships conditioned on, or accounting for, all other variables in the model, whereas traditional approaches may not control for other variables (34,36,39). These joint relationships form patterns of components by food groups and allow for an exploration of the internal structure of dietary patterns (39). An added benefit of this modelling approach is that results are presented using interpretable graphical representations such as networks, easing knowledge mobilization with diverse stakeholders, including policymakers.

Probabilistic graphical models can be extended to incorporate categorical variables, enabling explorations of joint relationships among mixed variable types (34,41). A growing number of studies have used mixed graphical models to jointly model dietary components in relation to other characteristics (32,33,42). For example, Hoang et al. (32) applied mixed graphical models to explore joint relationships among 16 dietary components, demographic characteristics, and health parameters. When dietary components were condensed to a single dietary score, no relationships with demographic or health characteristics were observed. However, when components were considered separately, joint relationships were observed between dietary components and other characteristics (32).

Given growing recognition of the need to account for dietary heterogeneity among populations (43,44), it is particularly relevant to explore the joint relationships between intersecting sociodemographic characteristics and high-dimensional dietary patterns. The objective of this study was to explore dietary patterns and their joint associations with sociodemographic characteristics among adults in Canada using mixed graphical models.

## Methods

### Data source and analytic sample

Data were drawn from the Public Use Microdata Files for the 2015 Canadian Community Health Survey (CCHS) Nutrition, a cross-sectional survey which contains information on a range of sociodemographic characteristics and up to two 24-hour dietary recalls (24HDRs) per participant (first recall n=20 487, second recall n=7623). Information about sociodemographic characteristics were collected during the first interview when participants completed the first recall. The 2015 CCHS Nutrition was conducted by Statistics Canada and Health Canada from January to December 2015 and is representative of individuals living in the 10 provinces in Canada (45). The survey included individuals aged one year and older who lived in private dwellings, with sampling based on age, sex, geographic area, and socioeconomic status (45). Individuals who lived in the territories, who were full-time members of the Canadian Forces, those living on reserves and in some remote areas, and those living in institutions were not included (45). Because publicly available data were used, Ethical Standards Disclosure was not applicable. Reporting follows the *Strengthening the Reporting of Observational Studies in Epidemiology-Nutrition* guidelines (46).

The analytic sample (n=14 097) consisted of adults aged 18 years and older who completed the first 24HDR, excluding those ≤17 years because of evidence suggesting the dietary patterns of children and youth are distinct from those of adults (47). Those with zero energy intakes (n=11) and/or missing data on characteristics of interest including household food security status (n=55), income (n=7), employment status (n=22), education (n=94), and smoking status (n=29) were also excluded from analysis (total excluded n=178). A comparison of included and excluded participants revealed a similar distribution according to age and sex (data not shown).

#### Sociodemographic characteristics

Sociodemographic characteristics of interest were selected because they have been shown to be related to dietary intake, individually or in some combination, in analyses focused on individual dietary components or using diet quality indices (22,23,25,48–52). A directed acyclic graph was created to explore which variables to include and their hypothesized directions with dietary patterns (**Supplementary Figure 1**), while considering what was available for analysis in the public-use version of the 2015 CCHS Nutrition (53,54). Because information on gender identity was not available, biological sex was included as a binary variable, with responses including male or female. Age in years was included as a continuous variable. Total household income was expressed using a categorical variable based on reported income, with categories of $0-19,999, $20,000-$39,999, $40,000-59,999, 60,000-79,999, $80,000-99,999, $100,000-119,000, $120,000-139,999, and $140,000 and higher. Household food security status was expressed using a categorical variable, determined based on affirmative responses to the Household Food Security Survey Module (55) and coded using Health Canada’s scoring thresholds. Categorizations of food security status included living in households that were food secure, moderately food insecure, or severely food insecure (56). Respondent education was included as a four-level categorical variable, with categorizations of less than a high school diploma or its equivalent; a high school diploma or its equivalent; a certificate or diploma at the trade, college, or below bachelor’s level; and a bachelor’s degree or higher. Employment status over the past week was condensed from a four-level variable to a binary variable reflecting those who did or did not work in the previous week. Geographic regions included the Maritimes, Québec, Ontario, the Prairies, and British Columbia. Smoking status was assessed as whether participants currently smoked cigarettes daily, occasionally, or not at all.

#### 24-hour dietary recalls

Dietary intake data were collected by trained interviewers using the Automated Multiple-Pass Method, originally designed by the United States Department of Agriculture (57–59) and adapted to the Canadian context by Health Canada (45). The Automated Multiple-Pass Method probes for commonly forgotten foods and beverages to minimize misreporting. Data from the first 24HDR for each participant were used. Foods and beverages reported were assigned food codes using the 2015 Canadian Nutrient File and a recipe file (45,60) and categorized into 156 food groups—referred to as the Bureau of Nutritional Sciences food groups—by Health Canada (61). Consistent with similar research (32,37), the number of food groups was reduced to 30 for analysis by condensing groups for which there was a high frequency of zero intakes and that included similar foods (e.g., potato chips, popcorn, and other salty snacks were grouped into a salty snack group) (**Supplementary Table 1**). Food group variables were expressed in grams consumed. To account for the skewed distribution typically observed in nutrition data and to better approximate normality, a log-transformation was applied to dietary variables, adding a constant of one to allow for the transformation of zero values (62,63).

### Statistical Analysis

#### Network analysis

Analyses were conducted using R and RStudio (RStudio, Boston, MA, 2020) (64). Mixed graphical models can be used to examine interrelationships among continuous and categorical variables, simultaneously modelling variables that follow a variety of distributions, including Gaussian and multinomial (41,65). The models used linear regression for Gaussian variables (dietary intake variables and age) and penalized multinomial logistic regression for categorical variables (the remaining sociodemographic variables) (34,41). Variables, or nodes, are connected by edges, which represent dependencies between nodes, conditioned on all other variables in the network (35,36). The absence of an edge represents conditional independence between nodes, conditioned on all other variables in the network (35,36). Graphically, the width of edges is proportionate to the strength of the edge parameter (34). Directionality can be assessed between continuous nodes, where positive or negative linear relationships are inferred, but the direction of relationships is undefined for relationships involving a categorical node (34). The ‘mgm’ package was used to model dependencies (34,66).

To reduce the possibility of overfitting and improve interpretability, a ‘least absolute shrinkage and selection operator’ (LASSO) regularisation parameter was used to perform variable selection and generate a sparse network by allowing some coefficients to be exactly equal to zero (66–68). To further improve interpretation of networks by not displaying very weak non-zero relationships, edges with weights below |0.05| were not shown (69). Ten-fold cross-validation was applied to determine the regularization parameter. To assess the stability of the network, bootstrapping with 2500 replication weights was used with the ‘bootnet’ package (70). The standard (root) estimates and bootstrapped confidence intervals are presented. Mixed graphical models were visualized using the ‘qgraph’ package based on the standard estimates (69).

Variables in the networks included the 30 food groups and the sociodemographic characteristics of interest. Three networks were modelled: network one included sociodemographic variables only, network two included dietary components only, and network three included both the dietary components and sociodemographic characteristics to explore their joint relationships. To facilitate interpretation in visualizing the networks, variables were assigned colour-coded group-level categories that included sociodemographic characteristics, fruit and vegetables, grains, protein foods, foods to limit, and other foods/beverages.

Network centrality indices were applied to indicate the importance of nodes in the network. Centrality was captured by three measures, including strength, which measured how much nodes were directly connected to other nodes; closeness, which measured how much nodes were indirectly connected to other nodes; and betweenness, which measured the importance of a node in the path of other nodes (71). Network stability was measured using the centrality stability coefficient, which indicates whether the ranking of nodes remains consistent when cases are dropped through bootstrapping (71). The centrality stability coefficient should be above 0.25, and ideally above 0.5 (71).

Survey weights from the 2015 CCHS Nutrition were incorporated to improve population representativeness (45).

## Results

### Sample characteristics

Characteristics of the sample are presented in **Table 1**. The mean age was 48.5 (SD ± 17.5) years and half of participants identified as female. About a quarter (28.1%) of participants had a bachelor’s degree or higher and 60.1% worked in the previous week. Mean one-day intakes of food groups are presented in **Table 2**.

**Table 1:** Characteristics of adults in the 2015 Canadian Community Health Survey Nutrition.

| **Characteristic** | **Total**<br>**(n=14,097)**<br>**Weighted %** |
| --- | --- |
| Sex |  |
| Male | 49.2 |
| Female | 50.8 |
| Age (mean, years) ± SD | 48.5 (17.5) |
| Household food security status |  |
| Food secure | 92.2 |
| Moderately food insecure | 5.3 |
| Severely food insecure | 2.5 |
| Income (household) |  |
| \$0-19,999 | 8.0 |
| \$20,000-39,999 | 16.7 |
| \$40,000-59,999 | 16.4 |
| \$60,000-79,999 | 14.1 |
| \$80,000-99,999 | 11.1 |
| \$100,000-119,999 | 10.3 |
| \$120,000-139,999 | 7.0 |
| \$>140,000 | 16.5 |
| Education (respondent) |  |
| Less than high school | 11.7 |
| High school diploma | 26.7 |
| Certificate/diploma below bachelor's | 33.6 |
| Bachelor's degree or higher | 28.1 |
| Worked in the past week |  |
| Yes | 60.1 |
| No | 39.9 |
| Geographic region |  |
| Maritimes | 6.8 |
| Québec | 23.5 |
| Ontario | 38.9 |
| Prairies | 17.7 |
| British Columbia | 13.1 |
| Smoking status |  |
| Daily | 13.7 |
| Occasionally | 4.4 |
| Not at all | 81.9 |

**Table 2:** Mean one-day intakes of 30 food groupings used to identify dietary pattern networks among adults in the 2015 Canadian Community Health Survey Nutrition.

| Food grouping* | Mean (grams) ± SD | Median (IQR)<sup>§</sup> |
| --- | --- | --- |
| Alcohol | 141 ± 380.8 | 0 (0-27.8) |
| Coffee & tea | 449.7 ± 454.1 | 380.4 (125.9-616.9) |
| Cured meats | 22.9 ± 51.7 | 0 (0-25.2) |
| Higher fat dairy products | 60.2 ± 121 | 17.8 (0-63) |
| Lower fat dairy products | 157.8 ± 215.6 | 64 (14.3-244) |
| Eggs | 25.4 ± 47.9 | 2.3 (0-37.4) |
| Fish and shellfish | 19 ± 55.3 | 0 (0-0) |
| Other grains (refined or whole grain) | 91.5 ± 119.3 | 44.1 (0-142.9) |
| Green vegetables | 41.5 ± 70.8 | 11.3 (0-55.6) |
| Onions, garlic, leeks, green onion | 14.2 ± 24.4 | 3.8 (0-18.8) |
| Orange vegetables | 16.2 ± 49.5 | 0 (0-10) |
| Other foods/beverages | 76 ± 188.2 | 16.3 (3.1-54.2) |
| Fruits (other) | 97 ± 155.7 | 16.4 (0-174.2) |
| Vegetables (other) | 47.9 ± 82.4 | 14.1 (0-61.3) |
| Plant-based proteins/alternatives | 35.2 ± 80.8 | 0 (0-32.4) |
| Potatoes | 42 ± 89.1 | 0 (0-50) |
| Poultry | 45.5 ± 86.9 | 0 (0-66.8) |
| Red meat | 45.2 ± 77.9 | 0 (0-70.2) |
| Refined grains | 63.5 ± 78.9 | 42 (0-99) |
| Salty snacks | 8.9 ± 29.7 | 0 (0-0) |
| Saturated fats<sup>a</sup> | 7.5 ± 13.2 | 2.1 (0-10) |
| Soups | 44.3 ± 115.4 | 0 (0-1.2) |
| Sugars<sup>b</sup> | 17.6 ± 28.2 | 8.3 (0-22.9) |
| Sweet beverages | 178.5 ± 310.3 | 0 (0-284.2) |
| Sweet treats/desserts | 32.7 ± 69.6 | 0 (0-35) |
| Tomatoes | 44.5 ± 77.8 | 2 (0-61.2) |
| Tropical fruits | 53.6 ± 87 | 0 (0-118) |
| Unsaturated fats | 10.3 ± 13.3 | 6.7 (1.2-14.5) |
| Water | 987.1 ± 902.6 | 781.3 (383.8-1402.5) |
| Whole grains | 40.9 ± 74.5 | 0 (0-64) |

\*Further details on food groupings are provided in Supplementary Table 1
<sup>§</sup>Median values provide an indication of the episodicity of food groupings
<sup>a</sup>Represents food sources of saturated fat (i.e., butter)
<sup>b</sup>Represents food sources of sugars (i.e., jams/jellies/marmalade)

#### Sociodemographic characteristic network

The first network considered only the sociodemographic characteristics (**Figure 1**). Sex shared an edge with household food security status (0.15; 95% CI [0.05, 0.25]), working in the past week (0.20; 95% CI [0.12, 0.29]) and smoking status (0.14; 95% CI [0.07, 0.22]) and was weakly related with other variables (<0.1). Geographic region shared edges with household food security (0.14; 95% CI [0.04, 0.24]), income (0.12; 95% CI [0.04, 0.20]), and education (0.16; 95% CI [0.06, 0.25]) and was weakly associated with other variables (<0.1). The strongest pairwise relationships were observed between household food security status and income (1.10; 95% CI [0.86, 1.32]), income and working in the past week (0.67; 95% CI [0.54, 0.80]), and age and working in the past week (0.53; 95% CI [0.47, 0.59]). Income shared edges with age (0.17; 95% CI [0.10, 0.23]) and education (0.40; 95% CI [0.28, 0.50]). Household food security status also shared edges with age (0.34; 95% CI [0.26, 0.43]), smoking status (0.37; 95% CI [0.23, 0.51]), education (0.14; 95% CI [0.02, 0.27]) and working in the past week (0.11; 95% CI [0.00, 0.22]). Further, working in the past week shared an edge with education (0.38; 95% CI [0.27, 0.50]) and smoking status (0.14; 95% CI [0.05, 0.24]). Smoking status additionally shared an edge with education (0.29; 95% CI [0.17, 0.42]) and age (0.19; 95% CI [0.12, 0.26]). Age also shared an edge with education (0.28; 95% CI [0.28, 0.35]). The weighted adjacency matrix describing all edge weights for the sociodemographic characteristic network is presented in **Supplemental Table 2**.

**Figure 1.**
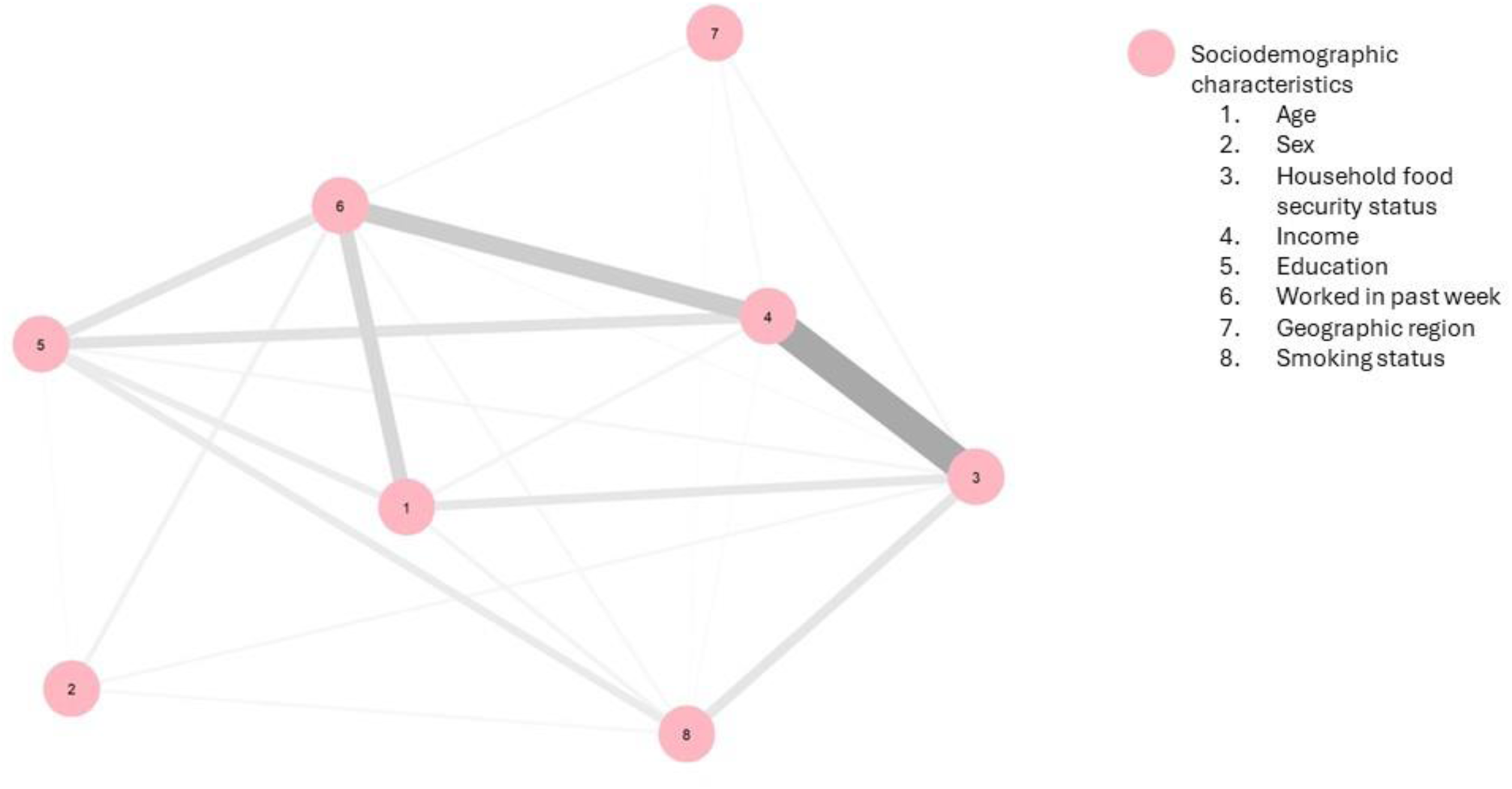
Network of sociodemographic characteristics among adults ≥18 years in the 2015 Canadian Community Health Survey Nutrition Network illustrates the distribution of nodes conditional on all other nodes in the network. Grey edges indicate an undefined relationship and are shown when categorical variables are involved in the relationship (34). Numbers assigned to variables do not indicate a ranking.

#### Dietary pattern network

The second network explored the joint relationships among dietary components (**Figure 2**). Several fruit and vegetables dietary components shared edges, with a positive linear relationship identified between tomatoes and onions and garlic (0.19; 95% CI [0.16, 0.23]). In turn, a positive linear relationship was observed between onions and garlic and both soup (0.24; 95% CI [0.21, 0.28]) and other vegetables (0.23; 95% CI [0.20, 0.27]). Soup and other vegetables were each positively associated with orange vegetables (0.17; 95% CI [0.14, 0.21], 0.13; 95% CI [0.09, 0.16] respectively). Orange vegetables and other vegetables were positively associated with green vegetables (0.12; 95% CI [0.08, 0.16], 0.10; 95% CI [0.07, 0.14] respectively). Tropical fruits and other fruits were also positively related (0.14; 95% CI [0.10, 0.18]).

**Figure 2.**
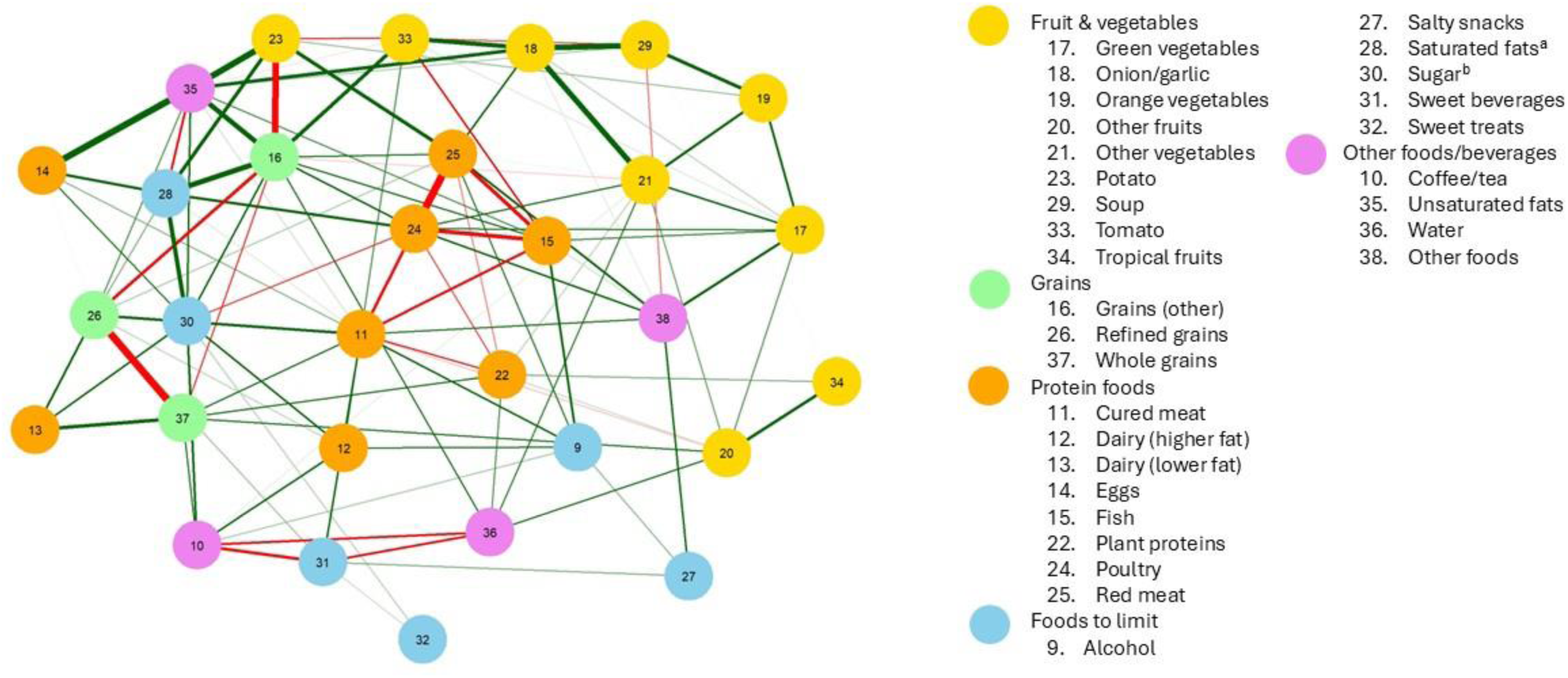
Network of dietary components among adults ≥18 years in the 2015 Canadian Community Health Survey Nutrition. Network illustrates the distribution of nodes conditional on all other nodes in the network. Green edges within the network represent positive associations between continuous variables, and red edges represent negative interactions between continuous variables (34). Numbers assigned to variables do not indicate a ranking. ^a^Represents food sources of saturated fat (i.e., butter) ^b^Represents food sources of sugars (i.e., jams/jellies/marmalade)

Negative linear relationships were identified among other components. A network of negative linear relationships was identified between cured meat and fish (−0.13; 95% CI [−0.16, −0.10]) cured meat and poultry (−0.14; 95% CI [−0.17, −0.11]), poultry and fish (−0.18; 95% CI [−0.21, −0.14]), poultry and red meat (−0.29; 95% CI [−0.33, −0.26]), and fish and red meat (−0.16; 95% CI [−0.20, −0.12]). Refined grains and whole grains were also negatively related (−0.30; 95% CI [−0.33, −0.26]). Negative linear relationships were observed between coffee/tea and water (−0.12; 95% CI [−0.16, −0.09]), coffee/tea and sweet beverages (−0.14; 95% CI [−0.18, −0.11]), and water and sweet beverages (−0.12; 95% CI [−0.16, −0.09]).

Patterns were also observed outside of groupings of similar foods. Eggs were positively associated with unsaturated fats (0.26; 95% CI [0.23, 0.30]), and unsaturated fats were positively associated with potatoes (0.25; 95% CI [0.21, 0.28]) and grains (other) (0.21; 95% CI [0.18, 0.25]). Grains (other) and potatoes were negatively associated (−0.27; 95% CI [−0.30, −0.23]) and grains (other) and saturated fats were positively associated (0.22; 95% CI [0.19, 0.25]). A positive linear relationship was observed between food sources of saturated fats and sugars (0.19; 95% CI [0.15, 0.22]). Weak relationships <0.1 were observed among other sweet food groups (sugars, sweet beverages, sweet treats). The weighted adjacency matrix describing all edge weights for the dietary component network is presented in **Supplemental Table 3**.

#### Dietary pattern and sociodemographic characteristic network

The third network included both the food group and sociodemographic variables (**Figure 3**). Accounting for the dietary components, sociodemographic characteristics largely patterned together, with similar pairwise relationships as observed in the network that explored only sociodemographic characteristics (Figure 1). Dietary components also maintained similar joint relationships as observed in the second network (Figure 2). For example, refined grains and whole grains were negatively linearly related (−0.29; 95% CI [−0.32, −0.25]) and the pattern observed among vegetable components was unchanged when accounting for sociodemographic variables. Sociodemographic characteristics were associated with several dietary components. Age was negatively linearly related with grains (other) (−0.12; 95% CI [−0.16, −0.09]) and positively related with coffee/tea (0.21; 95% CI [0.17, 0.24]) and whole grains (0.12; 95% CI [0.08, 0.15]). Sex shared edges with sweet beverages (0.11; 95% CI [0.06, 0.17]), alcohol (0.18; 95% CI [0.13, 0.24]), cured meat (0.20; 95% CI [0.15, 0.26]), and red meat (0.16; 95% CI [0.11, 0.21]). Smoking status shared an edge with whole grains (0.11; 95% CI [0.06, 0.17]) and education shared an edge with alcohol (0.10; 95% CI [0.04, 0.17]). Weak pairwise relationships were observed between other sociodemographic characteristics and dietary components. The weighted adjacency matrix describing all edge weights for the network of sociodemographic characteristics and dietary components is presented in **Supplemental Table 4**.

**Figure 3.**
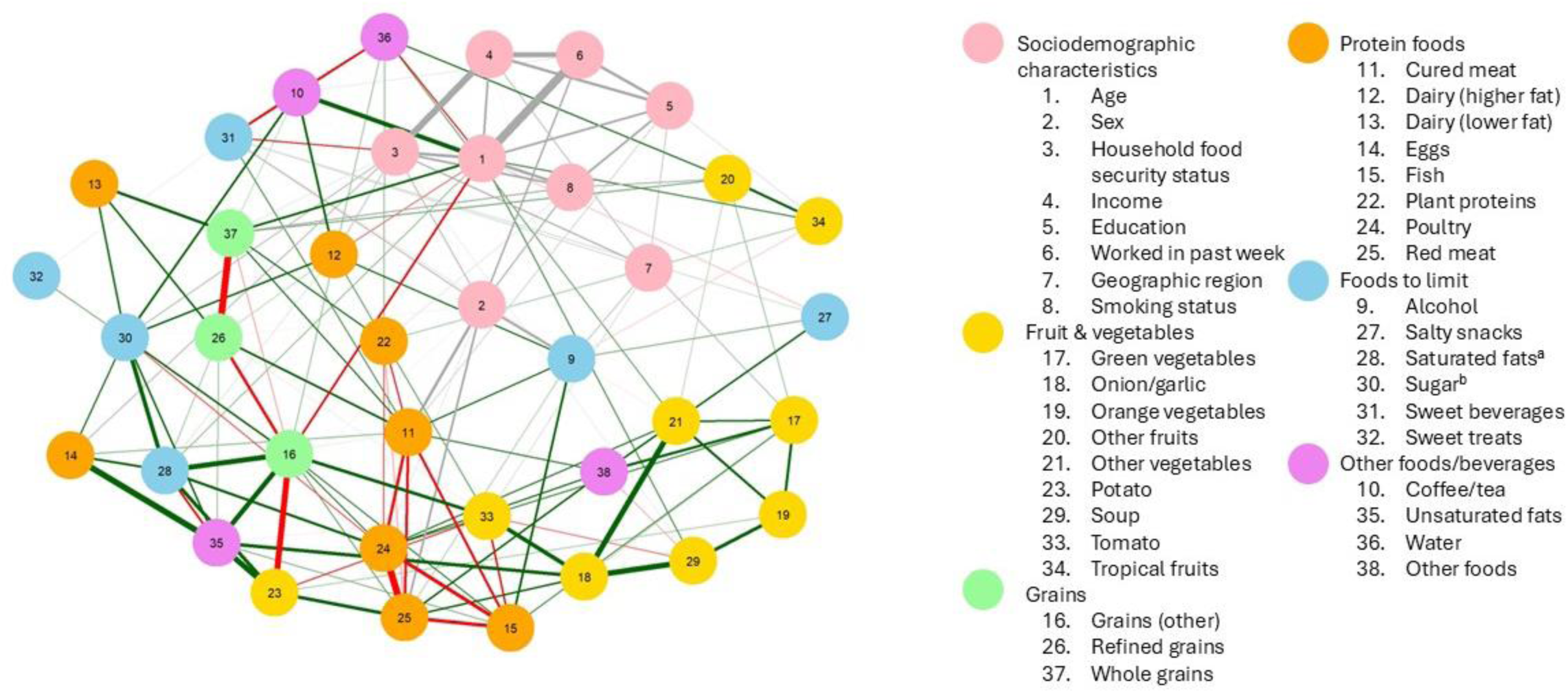
Network of dietary components and sociodemographic characteristics among adults ≥18 years in the 2015 Canadian Community Health Survey Nutrition. Network illustrates the distribution of nodes conditional on all other nodes in the network. Green edges within the network represent positive associations between continuous variables, red edges represent negative interactions between continuous variables, and grey edges indicate an undefined relationship and are shown when categorical variables are involved in the relationship (34). Numbers assigned to variables do not indicate a ranking. ^a^Represents food sources of saturated fat (i.e., butter) ^b^Represents food sources of sugars (i.e., jams/jellies/marmalade)

Network centrality indices indicating the importance of nodes for the full network that included sociodemographic characteristics and dietary components are presented in **Figure 4**. Household food security and age were the strongest nodes in the network, followed by income and working in the past week. Nodes with less influence were salty snacks, sweet treats, and dairy (lower fat).

**Figure 4.**
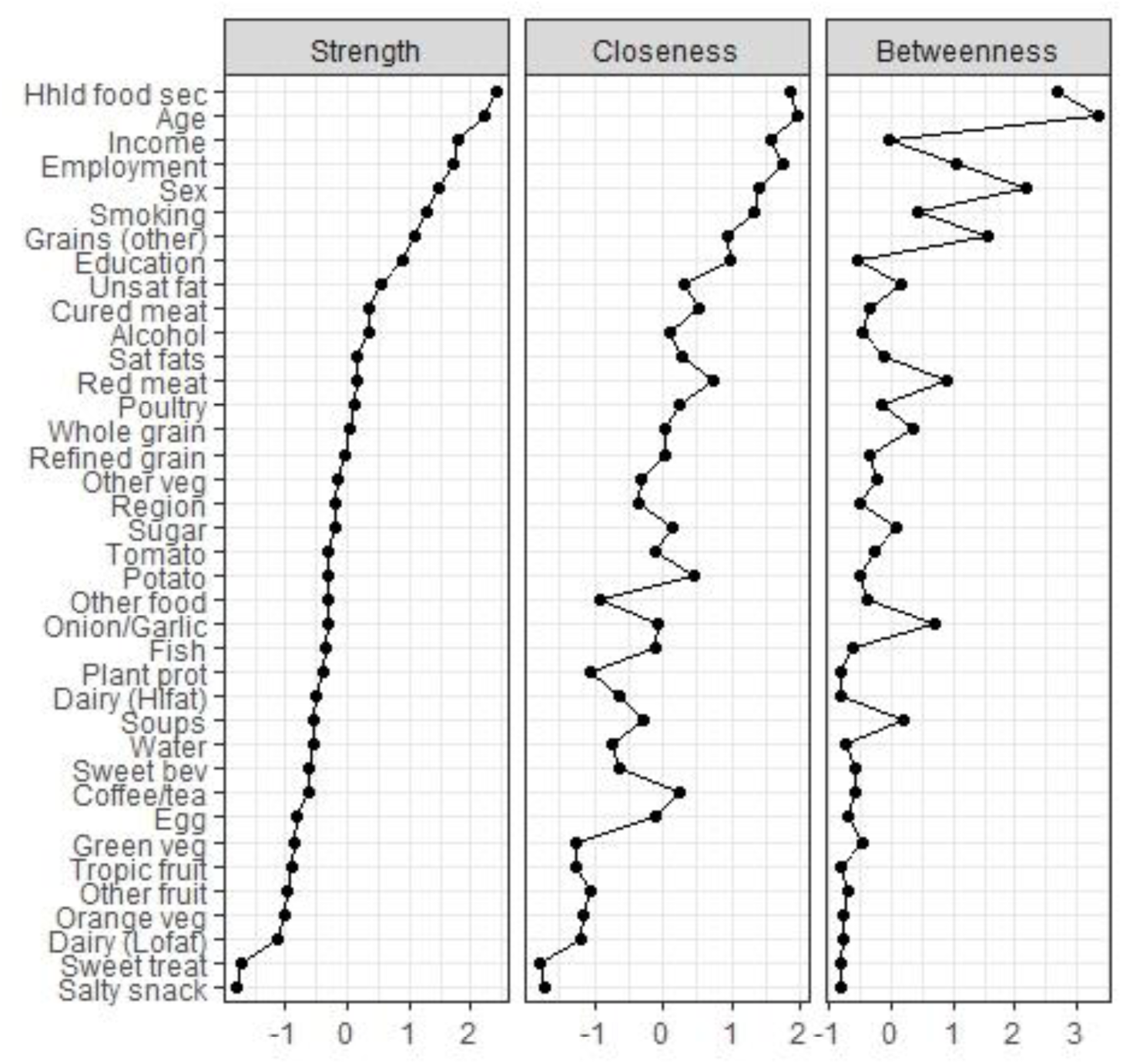
Centrality indices of variables included in the full network with sociodemographic and dietary components among adults ≥18 in the 2015 Canadian Community Health Survey Nutrition. Z scores are presented on the x axis, nodes are ordered by strength. Higher centrality indices indicate a greater importance of the node within the network.

The centrality stability coefficient was >0.5 for all three networks, indicating networks were suitably stable (71). Bootstrapped confidence intervals (CI) of estimated edge-weights for the full network are presented in **Supplementary Figure 2**.

## Discussion

This study explored two complex interrelated constructs—dietary patterns and intersecting sociodemographic characteristics—among adults in Canada. Mixed graphical models considered multidimensional relationships among dietary components and sociodemographic characteristics by conditioning each relationship between nodes on all other nodes in the network (34). The strongest pairwise relationships were observed among dietary components and among sociodemographic characteristics, rather than between dietary components and sociodemographic characteristics. For example, a strong edge was observed between income and household food security status, in line with associations observed through traditional approaches (72). The key sociodemographic characteristics that had pairwise relationships with dietary components were age and sex.

Similar to Iqbal et al. (39) in their work using separate Gaussian graphical models for males and females in a German adult population, we identified positive linear relationships among vegetable groupings. We also identified a comparable negative linear relationship between whole grains and refined grains, which was observed in the networks for both males and females in the German study (39). The observed negative linear relationship suggests that whole grains and refined grains displace each other within the dietary pattern after accounting for all other dietary components in the network. This finding is supportive of Canadian dietary guidance to emphasize whole grains over refined grains (73). In contrast with prior research, we observed negative linear relationships among several protein foods, whereas Iqbal et al. (39) observed positive partial correlations between processed meat and red meat, and red meat and poultry. These differences may be related in part to cross-country differences in dietary patterns between Canada and Germany and support the need for context-specific monitoring of dietary patterns. The negative linear relationships observed among protein foods in this study suggest that individuals may rely on a primary protein food type, regardless of whether the protein food is recommended (e.g., fish, poultry) or suggested to limit (e.g., cured meat) in dietary guidance, potentially highlighting an opportunity to improve messaging within such guidance.

Negative linear relationships were also observed among each of the beverage groups examined in this study, suggesting potential displacement, or prioritization, of beverage type. The 2019 Canada’s Food Guide recommends replacing sugary drinks with water (73), aligning with the observed negative relationship between consumption of water and consumption of other beverages, including sweet beverages and coffee. The positive linear relationship observed between food sources of sugars and saturated fats suggests these dietary components are consumed in combination, also relevant to guidance in the 2019 Canada’s Food Guide to limit intake of highly processed foods that add excess sugars, sodium, or saturated fats to the diets of Canadians (73). However, the foods captured in the relevant food groups in this study are considered to be processed culinary ingredients (e.g., sugar, lard) according to the NOVA classification of the degree of food processing (49).

Overall, our findings align with those of studies that have included sociodemographic characteristics and dietary components in mixed graphical models (32,33). In agreement with the current findings, the strongest relationships identified by Hoang et al. (32) were within dietary component groups and within sociodemographic characteristic groups, rather than between sociodemographic characteristics and dietary components. Slurink et al. (33) included a range of sociodemographic characteristics, dietary components, and health indicators in mixed graphical models and found several pairwise relationships among sociodemographic characteristics and several relationships among dietary components but relatively few relationships between sociodemographic characteristics and dietary components. Sex similarly had the strongest pairwise relationships with dietary components (33). In the current study, the strongest nodes in the network were household food security, age, employment and income, which are all tied to financial resources, and these variables were strongly related with each other.

It is possible that the simultaneous inclusion of multiple sociodemographic characteristics in networks may mask relationships with dietary components because the model conditions each relationship on all other variables. It may be useful in future work to use stratification to explore how dietary patterns described using Gaussian graphical models differ by sociodemographic characteristics. Identifying how sociodemographic characteristics intersect first and then exploring how these intersecting characteristics are related with dietary patterns may yield deeper insights into how individual characteristics jointly shape intake. Other types of network methods, such as mutual information, exponential random graphs, and neural networks, have also been applied to characterize dietary patterns and to assess intersecting relationships between sociodemographic characteristics (26,74–76) and may be worth considering for future explorations.

Some findings on relationships between dietary intake and sociodemographic characteristics are comparable with those from prior Canadian research that used traditional statistical approaches (21–23,77). For example, Brassard et al. (22) observed differences in scores on the Healthy Eating Food Index-2019, which assesses alignment of dietary intake with the 2019 Canada’s Food Guide healthy food choices recommendations, by sex, age, and smoking status. Likewise, we observed pairwise relationships between sex and age and several dietary components, which were in turn related to other dietary components. Evidence suggests dietary inequities in Canada in relation to household food security status, income, and education (21,23,25,48,50). For example, using data from CCHS Nutrition and multivariable linear regression, Olstad et al. (21) found that males and females with higher income and education had higher diet quality compared to those with lower income and education. In contrast, the current study found few relationships between income or education and dietary components. We observed weak relationships between household food security and several dietary components, with the strongest relationship with sweet beverages. However, strong relationships were observed between income and household food security and between income and working in the past week, consistent with the literature (72,78).

Although probabilistic graphical modelling is an *a posteriori* or data driven method rather than an *a priori* method and the algorithm itself is undirected/unsupervised, several stages in the process involved researcher decisions. The inclusion of an interdisciplinary research team, with expertise in dietary assessment, statistics, and network methods, helped to guide this decision-making process. Decisions included the selection of food groups and parameters to determine model sparsity. For example, food groupings could have been condensed further or differently, which may have changed the resulting dietary patterns and their associations with sociodemographic characteristics. We also could have applied Extended Bayesian Information Criterion (EBIC) to select the regularization parameter instead of cross-validation (34). Extended Bayesian Information Criterion is typically more conservative than cross-validation (34), but given our interest in learning potential relationships, a potentially less conservative regularization parameter was chosen to prioritize discovery and hypothesis generation. While networks hold appeal to address complexity in understanding dietary patterns, these models may not consistently outperform traditional approaches (79). However, as noted by Iqbal et al. (39), graphical models have the potential to complement current methods to expand our understanding of multidimensional relationships, though findings require confirmation using traditional analytic approaches.

As researchers look to novel analytic approaches, including probabilistic graphical models and machine learning methods, to answer questions of increasing complexity relevant to dietary patterns, they must consider trade-offs with interpretability (80). For example, neural networks have excellent predictive capabilities but have been criticized for being ‘black box’ models (80,81). Likely, a combination of traditional and novel analytic approaches will be needed to expand our understanding of dietary patterns, for example, by using novel methods to generate new hypotheses about how dietary components may be interrelated and traditional methods to test such hypotheses.

Strengths of this work were the use of a large, population-representative dataset with detailed dietary intake data and a range of sociodemographic variables, along with the involvement of an interdisciplinary team to guide the analysis and interpretation. Several caveats are also relevant to the interpretation of our findings. Although second recalls were available for a subset of the sample, we relied on data from the first recall and did not account for random within-person variation in intake or systematic error (82). These sources of error may have distorted the observed relationships (83). We considered a range of approaches to possibly mitigate the effects of the error, including incorporating energy intake in the model and including both available recalls. However, preliminary analysis (data not shown) showed that energy was jointly correlated with nearly all dietary components, making it challenging to decipher other relationships with dietary components. This was also observed in another study applying mixed graphical models (33). We were not able to include available first and second recalls as separate observations in our estimation, as this would violate independence assumptions (65). Future work could seek to tailor probabilistic graphical models to the nuances of dietary intake data, including its measurement error structure, potentially expanding the insights to be gleaned from these approaches (84). Such models will be applicable to many other exposures and outcomes within health that are measured with error.

Another possible limitation was the reliance on linear models, which may have missed possible complex non-linear relationships between variables of interest (34). Further, several key population groups were not included in the 2015 CCHS Nutrition, including those living in the territories and on reserves, limiting the representativeness of the findings (45). Although other characteristics, such as racial and ethnic identity are relevant to these analyses, we were limited by available characteristics in the Public Use Microdata Files. This study considered food groups, comprised of foods as they were eaten. However, future work could integrate key nutrients of interest, such as saturated fat, sugars, and sodium. In addition to sociodemographic characteristics, future analyses could also be extended to include health outcomes using directed graphical models.

## Conclusion

This research provides a lens into how dietary components are related with one another and with sociodemographic characteristics among adults in Canada. The findings overall align with those of studies using traditional approaches, supporting the potential of methods such as graphical networks to extend our understanding of multidimensional relationships among dietary components and other factors. Some observed interrelationships among dietary components are consistent with messaging in current Canadian dietary guidance that recognizes trade-offs, such as between water and sugary beverages. Additional research using such methods to examine how individuals with different characteristics consume foods and beverages in combination may inform future guidance that is appropriately tailored to different population groups. In sum, mixed graphical models appear to be a useful tool to complement existing methods of dietary pattern analysis, offering the potential to learn internal structures of dietary patterns and how they are related with other relevant variables of interest.

## Data Availability

The data used in this study are available through the Statistics Canada Data Liberation Initiative.

https://www150.statcan.gc.ca/n1/en/catalogue/82M0024X

## Acknowledgements

Statement of authors’ contributions: JMH designed the research, analyzed the data, drafted the manuscript, and had primary responsibility for final content. DS and AT-E provided guidance on the analytic approach. All authors read, provided feedback, and approved the final manuscript

The authors are grateful to the members of a working group that focused on the applicability of AI and related methods to expand understanding of dietary patterns, including Kevin W. Dodd, Amir Nazemi, Paul Fieguth, Jennifer Vena, Megan Deitchler, and Navreet Singh.

## Data sharing

The data used in this study are available through the Statistics Canada Data Liberation Initiative.

## Abbreviations

24HDR: 24-hour dietary recall
CCHS: Canadian Community Health Survey

