## Supplementary Materials for "Multidimensional dietary patterns and their joint associations with intersecting sociodemographic characteristics among adults in Canada: a cross-sectional study"

**Supplementary Figure 1:** Directed acyclic graph of relationships between sociodemographic characteristics of interest and dietary patterns

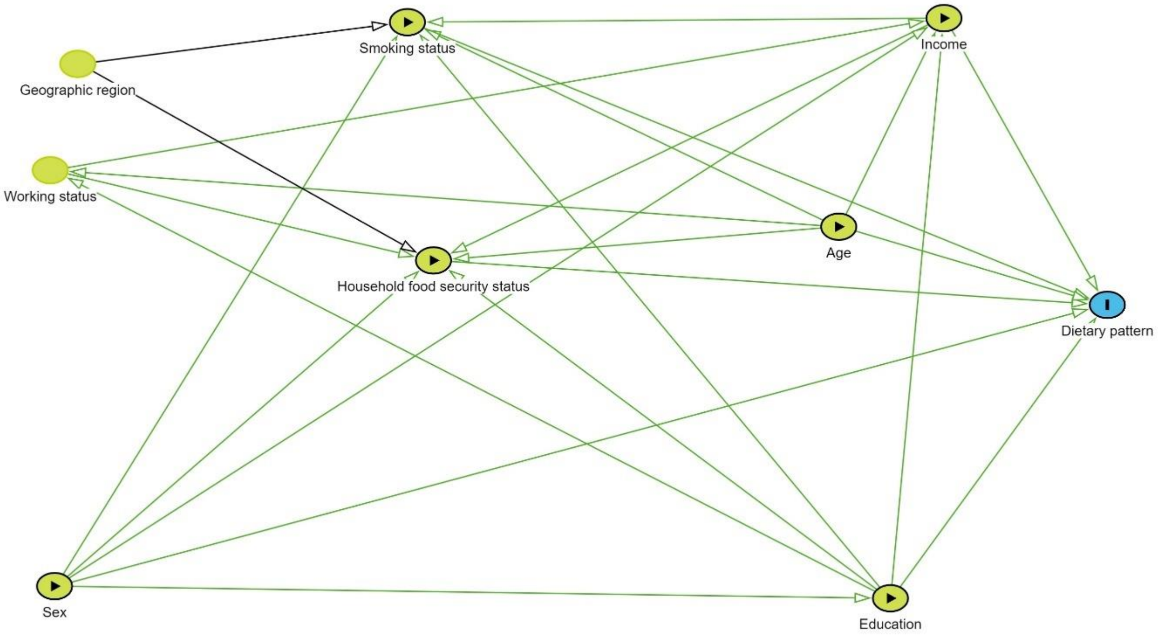

**Supplementary Figure 2:** Bootstrapped confidence intervals of edge weights for the full network including sociodemographic characteristics and dietary components among adults ≥18 y in the 2015 Canadian Community Health Survey Nutrition

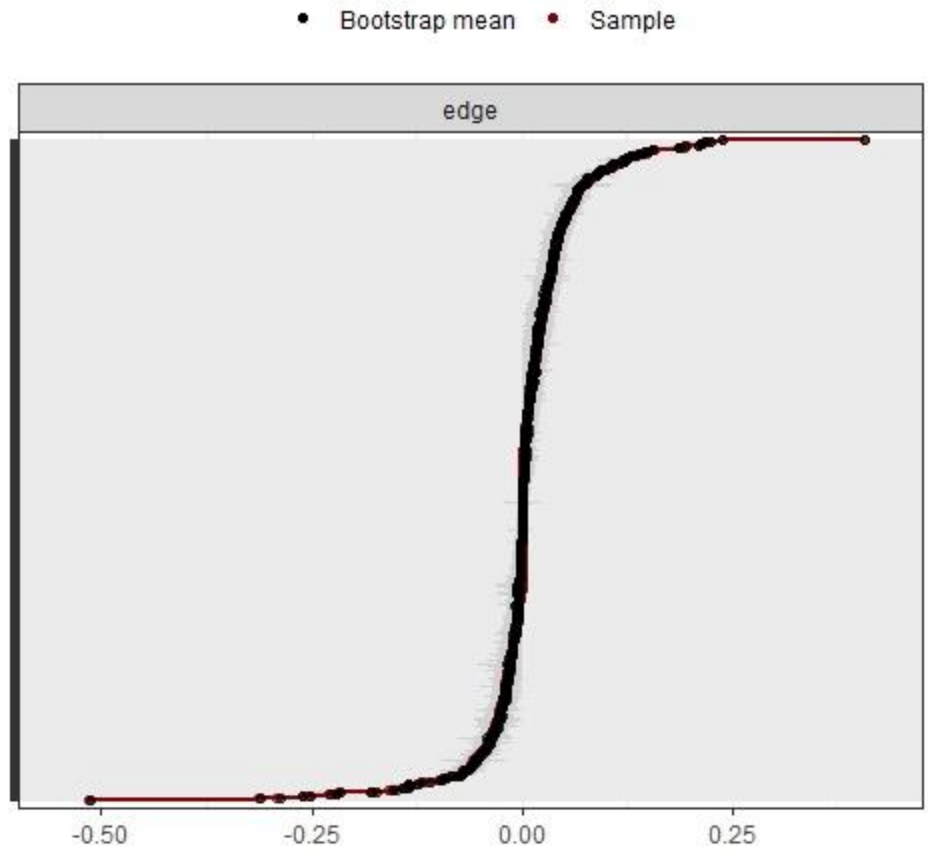

**Supplementary Table 1:** Bureau of Nutritional Science condensed variables for the analysis of dietary patterns in the 2015 CCHS Nutrition

| New variable | New variable description | % Zero intakes[1]: condensed variable | Original Variable | Description (gram weight) | % Zero intakes[1]: original variable |
|---|---|---|---|---|---|
| **Alcohol** | Alcohol | 74.9 | BNSD47A | Spirits | 95.4 |
| | | | BNSD47B | Liqueurs | 99.4 |
| | | | BNSD48A | Wine | 88.2 |
| | | | BNSD49A | Beer | 88.7 |
| | | | BNSD49B | Coolers | 100.0 |
| **CoffeeTea** | Coffee & tea | 17.8 | BNSD51A | Tea (Incl Iced Tea) | 65.6 |
| | | | BNSD51B | Coffee | 34.6 |
| **Curedmeat** | Cured meats | 62.8 | BNSD25C | Bacon | 89.7 |
| | | | BNSD25D | Ham: cured - lean only | 97.0 |
| | | | BNSD25E | Ham: cured - lean and fat | 98.1 |
| | | | BNSD30A | Sausage | 88.5 |
| | | | BNSD32A | Luncheon meat | 80.7 |
| **DairyHifat** | Higher fat dairy products | 31.0 | BNSD10A | Milk: whole | 73.8 |
| | | | BNSD10E | Milk: Evaporated whole | 98.6 |
| | | | BNSD10K | Milk: Goat/sheep | 99.9 |
| | | | BNSD13A | Whipping cream | 95.9 |
| | | | BNSD13B | Table cream | 91.3 |
| | | | BNSD13C | Half & Half cream | 89.2 |
| | | | BNSD13D | Sour cream | 94.6 |
| | | | BNSD14D | Cheese: More than 25% B.F. | 59.8 |
| | | | BNSD15B | Yoghurts: More than 2.1% B.F. | 87.5 |
| **DairyLofat** | | 15.9 | BNSD10B | Milk: 2% | 39.5 |

| | | | | | |
|---|---|---|---|---|---|
| | Lower fat dairy products | | BNSD10C | Milk: 1% | 68.5 |
| | | | BNSD10D | Milk: Skim | 69.2 |
| | | | BNSD10F | Milk: Evaporated 2% | 99.7 |
| | | | BNSD10G | Milk: Evaporated skim | 100.0 |
| | | | BNSD10H | Milk: Condensed | 99.7 |
| | | | BNSD10I | Milk: other (whey/buttermilk) | 99.2 |
| | | | BNSD14A | Cottage cheese | 98.1 |
| | | | BNSD14B | Cheese: less than 10% B.F. | 97.3 |
| | | | BNSD14C | Cheese: 10% B.F. to 25% B.F. | 69.5 |
| | | | BNSD15A | Yoghurts: Less than 2% B.F. | 92.5 |
| **Egg** | Eggs | 42.4 | BNSD16A | Egg | 42.4 |
| **Fish** | Fish and shellfish | 81.4 | BNSD34A | Fish: less than 6% total fat | 89.3 |
| | | | BNSD34B | Fish: superior or equal to 6% total fat | 91.4 |
| | | | BNSD35A | Shellfish | 94.5 |
| **Grainsother** | Other grains (refined or whole grain) | 27.0 | BNSD01A | Pasta | 79.6 |
| | | | BNSD01B | Rice | 77.0 |
| | | | BNSD01C | Cereal/grains/flours | 45.3 |
| **Greenveg** | Green vegetables | 42.2 | BNSD36A | Beans | 92.8 |
| | | | BNSD36B | Broccoli | 88.7 |
| | | | BNSD36C | Cabbage/Kale | 86.5 |
| | | | BNSD36H | Lettuce/Leafy greens (spinach/mustard greens) | 64.7 |
| | | | BNSD36K | Peas/snow peas | 89.5 |
| **Oniongar** | Onion, garlic, leeks, green onion | 41.5 | BNSD36J | Onion/green onion/leeks/garlic | 41.5 |
| **Orangeveg** | Orange vegetables | 65.7 | BNSD36E | Carrots | 68.8 |
| | | | BNSD36M | Squashes | 94.4 |
| **Other** | | 4.1 | BNSD16B | Egg substitutes | 99.9 |

| | | | | | |
|---|---|---|---|---|---|
| | Other foods/beverages | | BNSD41D | Sugar substitutes | 95.0 |
| | | | BNSD46B | Soft drink: diet | 92.0 |
| | | | BNSD52A | Babyfood products | 99.7 |
| | | | BNSD52B | Infant formula | 100.0 |
| | | | BNSD53B | Others (Baking soda/baking powder/yeast) | 59.0 |
| | | | BNSD54A | Energy bar | 99.6 |
| | | | BNSD54B | Protein bar/shake | 99.4 |
| | | | BNSD54C | Meal replacements | 96.0 |
| | | | BNSD99A | Mexican recipes | 100.0 |
| | | | BNSD36O | Juices: Tomato & vegetables | 97.4 |
| | | | BNSD50C | Gravies | 96.2 |
| | | | BNSD50D | Sauces (White/Bearnaise/Soya/Tartar/Ketchup) | 67.3 |
| | | | BNSD50E | Salad dressings (with or without oil) | 62.0 |
| | | | BNSD50F | Seasonings (salt/vinegar) | 14.8 |
| | | | BNSD53A | Spices | 71.1 |
| **Otherfruit** | Fruits (apples, cherries, grapes, melons, peaches, pears, plums, strawberries, etc.) | 43.7 | BNSD40B | Apple | 77.5 |
| | | | BNSD40D | Cherries | 97.1 |
| | | | BNSD40E | Grapes/raisins | 86.4 |
| | | | BNSD40F | Melons (canteloup/honeydew/watermelon) | 95.8 |
| | | | BNSD40G | Peaches/nectarines | 96.8 |
| | | | BNSD40H | Pears | 96.5 |
| | | | BNSD40J | Plums/prunes | 98.2 |
| | | | BNSD40K | Strawberries | 90.7 |

| | | | | | |
|---|---|---|---|---|---|
| | | | BNSD40L | Other fruits (blueberries/date/kiwi/fruit salad) | 72.6 |
| **Otherveg** | Vegetables (cauliflower, celery, corn, mushrooms, peppers, etc.) | 36.4 | BNSD36D | Cauliflower | 95.4 |
| | | | BNSD36F | Celery | 79.0 |
| | | | BNSD36G | Corn | 92.7 |
| | | | BNSD36I | Mushrooms | 87.8 |
| | | | BNSD36L | Peppers: red/green | 77.7 |
| | | | BNSD36P | Other veg (cucumber/beet/turnip) | 55.2 |
| **Plantproteins** | Plant-based proteins/alternatives (plant-based beverages, nuts, seeds, nut butters, legumes, tofu) | 56.3 | BNSD10J | Plant-based beverage (soy/almond/coconut) | 95.7 |
| | | | BNSD33A | Nuts | 78.5 |
| | | | BNSD33B | Seeds | 95.2 |
| | | | BNSD33C | Peanut butter/other nut spreads | 85.7 |
| | | | BNSD37A | Legume | 89.3 |
| | | | BNSD37B | Foods made with vegetable proteins (tofu) | 97.1 |
| **Potato** | Potatoes | 60.6 | BNSD38B | Fried/roasted potatoes | 88.9 |
| | | | BNSD39A | Potato | 69.8 |
| **Poultry** | Poultry | 60.9 | BNSD27A | Chicken: meat only | 70.0 |
| | | | BNSD27B | Chicken: meat and skin | 92.2 |
| | | | BNSD27C | Turkey: meat only | 97.4 |
| | | | BNSD27D | Turkey: meat + skin/ground | 98.8 |
| | | | BNSD27E | Other birds: Duck/pheasant/pigeon | 99.7 |
| | | | BNSD27F | Birds: skin only | 100.0 |
| **Redmeat** | Red meat | 54.6 | BNSD22A | Beef: Lean only | 89.8 |
| | | | BNSD22B | Beef: Lean + fat | 96.0 |
| | | | BNSD22C | Beef: ground | 78.1 |
| | | | BNSD23A | Veal: lean only | 99.7 |

| | | | | | |
|---|---|---|---|---|---|
| | | | BNSD23B | Veal: lean + fat/ground | 99.7 |
| | | | BNSD24A | Lamb: Lean only | 99.5 |
| | | | BNSD24B | Lamb: Lean + fat/ground | 99.7 |
| | | | BNSD25A | Pork: Fresh - lean only | 92.5 |
| | | | BNSD25B | Pork: Fresh - lean + fat/ground | 94.1 |
| | | | BNSD28A | Liver | 99.7 |
| | | | BNSD28B | Liver pate | 99.9 |
| | | | BNSD29A | Offal | 99.8 |
| | | | BNSD31A | Game meat | 99.0 |
| **Refinedgrain** | Refined grains (probable) | 30.9 | BNSD02A | White breads | 70.2 |
| | | | BNSD04A | Rolls/bagels/pita/croutons/dumplings/matzo/tortilla | 63.6 |
| | | | BNSD04B | Crackers/crispbreads | 88.5 |
| | | | BNSD04C | Muffins/English muffins | 97.2 |
| | | | BNSD04D | Pancakes/Waffles | 98.6 |
| | | | BNSD04E | Croissants/Piecrusts/Phyllo Dough | 98.3 |
| | | | BNSD04F | Dry Mixes (Cakes/Muffins/Pancakes) | 95.9 |
| | | | BNSD06A | Breakfast cereal (other) | 91.7 |
| **Saltysnack** | Salty snacks | 79.3 | BNSD38A | Potato chips | 90.2 |
| | | | BNSD42A | Plain popcorn/pretzels | 97.1 |
| | | | BNSD42B | Salty/high-fat snacks (incl tortilla chips) | 90.9 |
| **Satfats** | Saturated fats | 44.4 | BNSD17A | Butter | 71.3 |
| | | | BNSD20A | Block margarine | 99.4 |
| | | | BNSD21B | Animal fats | 94.7 |
| | | | BNSD21C | Shortening | 63.5 |
| **Soup** | Soups | 74.4 | BNSD50A | Soups with vegetables | 93.0 |
| | | | BNSD50B | Soups without vegetables | 80.3 |

| | | | | | |
|---|---|---|---|---|---|
| **Sugar** | Sugars (sugar, jams, syrup, honey, etc.) | 28.1 | BNSD41A | Sugars: white/brown | 36.4 |
| | | | BNSD41B | Jams/jellies/marmalade | 88.7 |
| | | | BNSD41C | Other sugars (syrups/molasses/honey) | 77.7 |
| **Sweetbev** | Sweet beverages | 50.6 | BNSD45A | Fruit juice | 70.0 |
| | | | BNSD46A | Soft drink: regular | 82.8 |
| | | | BNSD46C | Fruit drinks | 92.4 |
| | | | BNSD46D | Other beverages (malted milk/chocolate beverage) | 97.9 |
| | | | BNSD46E | Energy drink | 99.4 |
| | | | BNSD46F | Vitamin water | 99.6 |
| | | | BNSD46G | Sports drink | 98.7 |
| **Sweettreat** | Sweet treats/desserts | 50.7 | BNSD07A | Cookies (commercial) | 83.7 |
| | | | BNSD07B | Biscuits: commercial | 99.4 |
| | | | BNSD07C | Granola bar | 93.8 |
| | | | BNSD08A | Pies: commercial | 99.7 |
| | | | BNSD08B | Cakes: commercial (frozen cake) | 99.4 |
| | | | BNSD08C | Danishes/doughnuts/other pastries: commercial | 99.8 |
| | | | BNSD09A | Ice cream | 90.1 |
| | | | BNSD09B | Ice milk | 99.5 |
| | | | BNSD09C | Frozen yoghurt | 99.1 |
| | | | BNSD43A | Candy/gum | 90.9 |
| | | | BNSD43B | Ice pop/sherbert | 98.7 |
| | | | BNSD43C | Gelatin/dessert toppings/pudding mixes: commercial | 97.0 |
| | | | BNSD44A | Chocolate bar | 82.5 |
| **Tomato** | Tomatoes | 54.0 | BNSD36N | Tomatoes | 54.0 |

| | | | | | |
|---|---|---|---|---|---|
| **Tropicalfruit** | Tropical fruits (citrus, banana, pineapple) | 59.8 | BNSD40A | Citrus fruit (oranges/lemons/grapefruits) | 81.9 |
| | | | BNSD40C | Banana | 73.1 |
| | | | BNSD40I | Pineapple | 95.6 |
| **Unsatfat** | Unsaturated fats | 22.2 | BNSD18A | Regular margarine | 47.2 |
| | | | BNSD18B | Calorie-reduced margarine | 96.5 |
| | | | BNSD21A | Vegetable oils | 49.3 |
| **Water** | Water | 7.7 | BNSD51C | Water (Well/mineral) | 7.7 |
| **Wholegrain** | Whole grains | 52.8 | BNSD03A | Whole wheat breads | 74.5 |
| | | | BNSD03B | Other whole grain breads | 75.9 |
| | | | BNSD05A | Whole Grain/Oats/High Fibre Breakfast cereals | 75.2 |

[1] Day 1 intakes only

**Supplemental Table 2**: Weighted adjacency matrix network only sociodemographic characteristics among adults ≥18 years in the 2015 Canadian Community Health Survey Nutrition

| | **1** | **2** | **3** | **4** | **5** | **6** | **7** | **8** |
|---|---|---|---|---|---|---|---|---|
| **1** | 0.00 | 0.00 | 0.34 | 0.17 | 0.28 | 0.53 | 0.05 | 0.19 |
| **2** | 0.00 | 0.00 | 0.15 | 0.07 | 0.08 | 0.20 | 0.00 | 0.14 |
| **3** | 0.34 | 0.15 | 0.00 | 1.10 | 0.14 | 0.11 | 0.14 | 0.37 |
| **4** | 0.17 | 0.07 | 1.10 | 0.00 | 0.40 | 0.67 | 0.12 | 0.09 |
| **5** | 0.28 | 0.08 | 0.14 | 0.40 | 0.00 | 0.38 | 0.16 | 0.29 |
| **6** | 0.53 | 0.20 | 0.11 | 0.67 | 0.38 | 0.00 | 0.06 | 0.14 |
| **7** | 0.05 | 0.00 | 0.14 | 0.12 | 0.16 | 0.06 | 0.00 | 0.08 |
| **8** | 0.19 | 0.14 | 0.37 | 0.09 | 0.29 | 0.14 | 0.08 | 0.00 |

Legend: 1-Age, 2-Sex, 3-Household food security status, 4-Income, 5-Education, 6-Worked in the past week, 7-Geography, 8-Smoking status

**Supplemental Table 3**: Weighted adjacency matrix network only dietary components among adults ≥18 years in the 2015 Canadian Community Health Survey Nutrition

| | **1** | **2** | **3** | **4** | **5** | **6** | **7** | **8** | **9** | **10** | **11** | **12** | **13** | **14** | **15** |
|---|---|---|---|---|---|---|---|---|---|---|---|---|---|---|---|
| **1** | 0.00 | 0.06 | 0.10 | 0.09 | -0.02 | 0.02 | 0.12 | -0.03 | 0.00 | 0.03 | 0.00 | -0.02 | 0.05 | 0.00 | 0.04 |
| **2** | 0.06 | 0.00 | 0.01 | 0.10 | 0.00 | 0.05 | 0.02 | 0.00 | 0.02 | 0.00 | 0.00 | 0.00 | 0.05 | 0.03 | 0.00 |
| **3** | 0.10 | 0.01 | 0.00 | 0.03 | 0.03 | 0.06 | -0.13 | -0.03 | -0.01 | 0.00 | -0.04 | -0.06 | -0.02 | -0.09 | 0.00 |
| **4** | 0.09 | 0.10 | 0.03 | 0.00 | 0.02 | 0.00 | -0.04 | 0.00 | 0.02 | 0.00 | 0.00 | 0.02 | 0.04 | 0.03 | 0.00 |
| **5** | -0.02 | 0.00 | 0.03 | 0.02 | 0.00 | 0.02 | -0.01 | 0.03 | -0.01 | 0.00 | 0.01 | 0.03 | 0.01 | -0.05 | 0.03 |
| **6** | 0.02 | 0.05 | 0.06 | 0.00 | 0.02 | 0.00 | 0.00 | 0.00 | 0.02 | -0.01 | 0.00 | 0.01 | 0.00 | -0.03 | -0.03 |
| **7** | 0.12 | 0.02 | -0.13 | -0.04 | -0.01 | 0.00 | 0.00 | 0.08 | 0.07 | -0.02 | -0.01 | 0.02 | 0.02 | -0.05 | 0.00 |
| **8** | -0.03 | 0.00 | -0.03 | 0.00 | 0.03 | 0.00 | 0.08 | 0.00 | -0.02 | 0.04 | 0.04 | 0.00 | -0.06 | 0.00 | -0.27 |
| **9** | 0.00 | 0.02 | -0.01 | 0.02 | -0.01 | 0.02 | 0.07 | -0.02 | 0.00 | 0.06 | 0.12 | 0.07 | 0.10 | 0.02 | 0.00 |
| **10** | 0.03 | 0.00 | 0.00 | 0.00 | 0.00 | -0.01 | -0.02 | 0.04 | 0.06 | 0.00 | 0.04 | 0.02 | 0.23 | 0.05 | 0.02 |
| **11** | 0.00 | 0.00 | -0.04 | 0.00 | 0.01 | 0.00 | -0.01 | 0.04 | 0.12 | 0.04 | 0.00 | 0.04 | 0.13 | 0.04 | 0.06 |
| **12** | -0.02 | 0.00 | -0.06 | 0.02 | 0.03 | 0.01 | 0.02 | 0.00 | 0.07 | 0.02 | 0.04 | 0.00 | 0.07 | 0.06 | 0.00 |
| **13** | 0.05 | 0.05 | -0.02 | 0.04 | 0.01 | 0.00 | 0.02 | -0.06 | 0.10 | 0.23 | 0.13 | 0.07 | 0.00 | 0.06 | -0.03 |
| **14** | 0.00 | 0.03 | -0.09 | 0.03 | -0.05 | -0.03 | -0.05 | 0.00 | 0.02 | 0.05 | 0.04 | 0.06 | 0.06 | 0.00 | -0.03 |
| **15** | 0.04 | 0.00 | 0.00 | 0.00 | 0.03 | -0.03 | 0.00 | -0.27 | 0.00 | 0.02 | 0.06 | 0.00 | -0.03 | -0.03 | 0.00 |
| **16** | 0.01 | 0.00 | -0.14 | -0.02 | -0.02 | 0.00 | -0.18 | 0.09 | 0.08 | 0.00 | 0.02 | 0.00 | 0.08 | -0.07 | 0.02 |
| **17** | 0.07 | -0.01 | -0.12 | -0.02 | 0.00 | 0.00 | -0.16 | 0.08 | 0.01 | 0.10 | 0.00 | -0.02 | 0.05 | -0.06 | 0.16 |
| **18** | -0.04 | 0.03 | 0.12 | 0.06 | 0.11 | -0.05 | 0.00 | -0.15 | -0.02 | 0.01 | -0.04 | -0.01 | -0.01 | 0.02 | -0.01 |
| **19** | 0.06 | 0.02 | 0.04 | 0.03 | 0.00 | 0.00 | -0.02 | -0.02 | -0.01 | -0.02 | 0.00 | -0.04 | -0.02 | 0.00 | 0.00 |
| **20** | 0.03 | 0.00 | 0.05 | 0.05 | 0.04 | 0.12 | 0.00 | 0.22 | -0.04 | 0.03 | 0.00 | 0.00 | -0.01 | -0.04 | 0.17 |
| **21** | -0.02 | 0.02 | -0.03 | -0.04 | -0.01 | -0.03 | -0.03 | 0.01 | 0.01 | 0.24 | 0.17 | -0.03 | 0.00 | -0.03 | -0.03 |
| **22** | 0.00 | 0.12 | -0.03 | 0.09 | 0.10 | 0.08 | -0.02 | 0.10 | -0.04 | 0.00 | -0.01 | 0.04 | -0.03 | 0.02 | 0.01 |
| **23** | -0.05 | -0.14 | 0.09 | 0.03 | -0.01 | 0.04 | 0.01 | 0.03 | 0.00 | 0.02 | -0.01 | -0.03 | 0.01 | 0.00 | 0.03 |
| **24** | -0.04 | -0.02 | 0.00 | 0.04 | 0.03 | -0.04 | 0.00 | -0.01 | 0.00 | -0.04 | 0.00 | 0.00 | 0.02 | 0.04 | 0.00 |
| **25** | 0.03 | 0.00 | 0.02 | 0.07 | 0.03 | -0.03 | -0.11 | 0.18 | 0.05 | 0.19 | -0.01 | 0.02 | 0.05 | 0.00 | -0.08 |
| **26** | -0.04 | 0.02 | 0.01 | 0.02 | 0.03 | 0.02 | 0.03 | 0.00 | 0.04 | -0.01 | 0.03 | 0.14 | 0.02 | 0.06 | -0.01 |

| | | | | | | | | | | | | | | | |
|---|---|---|---|---|---|---|---|---|---|---|---|---|---|---|---|
| **27** | -0.01 | 0.00 | 0.05 | 0.02 | 0.05 | 0.26 | 0.07 | 0.21 | 0.00 | 0.15 | 0.05 | 0.00 | 0.03 | 0.03 | 0.25 |
| **28** | -0.02 | -0.12 | 0.01 | 0.01 | -0.02 | 0.02 | 0.01 | 0.08 | 0.03 | 0.01 | 0.01 | 0.08 | 0.07 | 0.07 | -0.02 |
| **29** | -0.02 | 0.08 | 0.09 | 0.00 | 0.16 | -0.02 | 0.02 | -0.07 | 0.00 | 0.00 | 0.00 | 0.08 | 0.03 | 0.08 | -0.02 |
| **30** | 0.00 | -0.02 | 0.08 | 0.05 | 0.03 | 0.03 | 0.05 | -0.02 | 0.12 | 0.05 | -0.03 | 0.00 | 0.04 | 0.02 | 0.03 |

| | **16** | **17** | **18** | **19** | **20** | **21** | **22** | **23** | **24** | **25** | **26** | **27** | **28** | **29** | **30** |
|---|---|---|---|---|---|---|---|---|---|---|---|---|---|---|---|
| **1** | 0.01 | 0.07 | -0.04 | 0.06 | 0.03 | -0.02 | 0.00 | -0.05 | -0.04 | 0.03 | -0.04 | -0.01 | -0.02 | -0.02 | 0.00 |
| **2** | 0.00 | -0.01 | 0.03 | 0.02 | 0.00 | 0.02 | 0.12 | -0.14 | -0.02 | 0.00 | 0.02 | 0.00 | -0.12 | 0.08 | -0.02 |
| **3** | -0.14 | -0.12 | 0.12 | 0.04 | 0.05 | -0.03 | -0.03 | 0.09 | 0.00 | 0.02 | 0.01 | 0.05 | 0.01 | 0.09 | 0.08 |
| **4** | -0.02 | -0.02 | 0.06 | 0.03 | 0.05 | -0.04 | 0.09 | 0.03 | 0.04 | 0.07 | 0.02 | 0.02 | 0.01 | 0.00 | 0.05 |
| **5** | -0.02 | 0.00 | 0.11 | 0.00 | 0.04 | -0.01 | 0.10 | -0.01 | 0.03 | 0.03 | 0.03 | 0.05 | -0.02 | 0.16 | 0.03 |
| **6** | 0.00 | 0.00 | -0.05 | 0.00 | 0.12 | -0.03 | 0.08 | 0.04 | -0.04 | -0.03 | 0.02 | 0.26 | 0.02 | -0.02 | 0.03 |
| **7** | -0.18 | -0.16 | 0.00 | -0.02 | 0.00 | -0.03 | -0.02 | 0.01 | 0.00 | -0.11 | 0.03 | 0.07 | 0.01 | 0.02 | 0.05 |
| **8** | 0.09 | 0.08 | -0.15 | -0.02 | 0.22 | 0.01 | 0.10 | 0.03 | -0.01 | 0.18 | 0.00 | 0.21 | 0.08 | -0.07 | -0.02 |
| **9** | 0.08 | 0.01 | -0.02 | -0.01 | -0.04 | 0.01 | -0.04 | 0.00 | 0.00 | 0.05 | 0.04 | 0.00 | 0.03 | 0.00 | 0.12 |
| **10** | 0.00 | 0.10 | 0.01 | -0.02 | 0.03 | 0.24 | 0.00 | 0.02 | -0.04 | 0.19 | -0.01 | 0.15 | 0.01 | 0.00 | 0.05 |
| **11** | 0.02 | 0.00 | -0.04 | 0.00 | 0.00 | 0.17 | -0.01 | -0.01 | 0.00 | -0.01 | 0.03 | 0.05 | 0.01 | 0.00 | -0.03 |
| **12** | 0.00 | -0.02 | -0.01 | -0.04 | 0.00 | -0.03 | 0.04 | -0.03 | 0.00 | 0.02 | 0.14 | 0.00 | 0.08 | 0.08 | 0.00 |
| **13** | 0.08 | 0.05 | -0.01 | -0.02 | -0.01 | 0.00 | -0.03 | 0.01 | 0.02 | 0.05 | 0.02 | 0.03 | 0.07 | 0.03 | 0.04 |
| **14** | -0.07 | -0.06 | 0.02 | 0.00 | -0.04 | -0.03 | 0.02 | 0.00 | 0.04 | 0.00 | 0.06 | 0.03 | 0.07 | 0.08 | 0.02 |
| **15** | 0.02 | 0.16 | -0.01 | 0.00 | 0.17 | -0.03 | 0.01 | 0.03 | 0.00 | -0.08 | -0.01 | 0.25 | -0.02 | -0.02 | 0.03 |
| **16** | 0.00 | -0.29 | 0.00 | 0.00 | 0.13 | 0.02 | -0.07 | 0.02 | 0.00 | -0.03 | -0.01 | 0.03 | 0.03 | -0.02 | 0.09 |
| **17** | -0.29 | 0.00 | 0.06 | 0.00 | -0.04 | -0.02 | 0.00 | 0.03 | 0.00 | 0.00 | -0.02 | 0.01 | 0.01 | -0.03 | 0.11 |
| **18** | 0.00 | 0.06 | 0.00 | 0.01 | 0.06 | 0.01 | 0.04 | 0.06 | 0.02 | 0.06 | -0.01 | 0.07 | 0.02 | -0.30 | 0.05 |
| **19** | 0.00 | 0.00 | 0.01 | 0.00 | 0.01 | 0.00 | 0.01 | 0.06 | 0.03 | 0.00 | 0.03 | -0.01 | 0.01 | -0.02 | 0.10 |
| **20** | 0.13 | -0.04 | 0.06 | 0.01 | 0.00 | 0.00 | 0.19 | 0.03 | 0.02 | -0.02 | 0.01 | -0.12 | 0.00 | 0.00 | 0.03 |
| **21** | 0.02 | -0.02 | 0.01 | 0.00 | 0.00 | 0.00 | 0.00 | -0.01 | 0.00 | -0.07 | 0.05 | 0.06 | -0.03 | 0.04 | -0.07 |
| **22** | -0.07 | 0.00 | 0.04 | 0.01 | 0.19 | 0.00 | 0.00 | 0.04 | 0.06 | -0.02 | 0.00 | 0.10 | 0.00 | -0.01 | 0.00 |
| **23** | 0.02 | 0.03 | 0.06 | 0.06 | 0.03 | -0.01 | 0.04 | 0.00 | 0.06 | 0.01 | -0.04 | 0.02 | -0.12 | -0.01 | -0.04 |

| | | | | | | | | | | | | | | | |
|---|---|---|---|---|---|---|---|---|---|---|---|---|---|---|---|
| **24** | 0.00 | 0.00 | 0.02 | 0.03 | 0.02 | 0.00 | 0.06 | 0.06 | 0.00 | 0.00 | 0.00 | 0.02 | 0.02 | 0.03 | 0.04 |
| **25** | -0.03 | 0.00 | 0.06 | 0.00 | -0.02 | -0.07 | -0.02 | 0.01 | 0.00 | 0.00 | 0.02 | -0.05 | 0.03 | 0.02 | -0.01 |
| **26** | -0.01 | -0.02 | -0.01 | 0.03 | 0.01 | 0.05 | 0.00 | -0.04 | 0.00 | 0.02 | 0.00 | 0.00 | 0.02 | 0.04 | -0.04 |
| **27** | 0.03 | 0.01 | 0.07 | -0.01 | -0.12 | 0.06 | 0.10 | 0.02 | 0.02 | -0.05 | 0.00 | 0.00 | 0.00 | -0.01 | 0.04 |
| **28** | 0.03 | 0.01 | 0.02 | 0.01 | 0.00 | -0.03 | 0.00 | -0.12 | 0.02 | 0.03 | 0.02 | 0.00 | 0.00 | 0.03 | 0.00 |
| **29** | -0.02 | -0.03 | -0.30 | -0.02 | 0.00 | 0.04 | -0.01 | -0.01 | 0.03 | 0.02 | 0.04 | -0.01 | 0.03 | 0.00 | 0.02 |
| **30** | 0.09 | 0.11 | 0.05 | 0.10 | 0.03 | -0.07 | 0.00 | -0.04 | 0.04 | -0.01 | -0.04 | 0.04 | 0.00 | 0.02 | 0.00 |

Legend: 1-Alcohol, 2-Coffee/tea, 3-Cured meat, 4-Dairy (higher fat), 5-Dairy (lower fat), 6-Eggs, 7-Fish, 8-Grains (other), 9-Green vegetables, 10-Onion/garlic, 11-Orange vegetables, 12-Other fruits, 13-Other vegetables, 14-Plant proteins, 15-Potato, 16-Poultry, 17-Red meat, 18-Refined grains, 19-Salty snacks, 20-Saturated fats, 21-Soup, 22-Sugar, 23-Sweet beverages, 24-Sweet treats, 25-Tomato, 26-Tropical fruits, 27-Unsaturated fats, 28-Water, 29-Whole grains, 30-Other foods

**Supplemental Table 4**: Weighted adjacency matrix of network including dietary components and sociodemographic characteristics among adults ≥18 years in the 2015 Canadian Community Health Survey Nutrition

| | **1** | **2** | **3** | **4** | **5** | **6** | **7** | **8** | **9** | **10** | **11** | **12** | **13** | **14** | **15** | **16** | **17** | **18** | **19** |
|---|---|---|---|---|---|---|---|---|---|---|---|---|---|---|---|---|---|---|---|
| **1** | 0.00 | 0.03 | 0.28 | 0.15 | 0.25 | 0.50 | 0.02 | 0.19 | 0.04 | 0.21 | -0.02 | -0.07 | 0.00 | 0.00 | 0.00 | -0.12 | 0.00 | 0.00 | 0.01 |
| **2** | 0.03 | 0.00 | 0.13 | 0.05 | 0.08 | 0.20 | 0.03 | 0.15 | 0.18 | 0.03 | 0.20 | 0.05 | 0.04 | 0.00 | 0.03 | 0.06 | 0.05 | 0.00 | 0.00 |
| **3** | 0.28 | 0.13 | 0.00 | 0.69 | 0.05 | 0.12 | 0.12 | 0.27 | 0.07 | 0.01 | 0.00 | 0.09 | 0.00 | 0.09 | 0.00 | 0.07 | 0.04 | 0.00 | 0.03 |
| **4** | 0.15 | 0.05 | 0.69 | 0.00 | 0.20 | 0.58 | 0.09 | 0.06 | 0.08 | 0.03 | 0.02 | 0.02 | 0.01 | 0.02 | 0.01 | 0.02 | 0.02 | 0.01 | 0.02 |
| **5** | 0.25 | 0.08 | 0.05 | 0.20 | 0.00 | 0.35 | 0.08 | 0.20 | 0.10 | 0.04 | 0.07 | 0.03 | 0.01 | 0.03 | 0.01 | 0.00 | 0.02 | 0.01 | 0.01 |
| **6** | 0.50 | 0.20 | 0.12 | 0.58 | 0.35 | 0.00 | 0.03 | 0.12 | 0.03 | 0.09 | 0.00 | 0.00 | 0.00 | 0.01 | 0.00 | 0.00 | 0.00 | 0.02 | 0.00 |
| **7** | 0.02 | 0.03 | 0.12 | 0.09 | 0.08 | 0.03 | 0.00 | 0.04 | 0.08 | 0.03 | 0.01 | 0.02 | 0.05 | 0.02 | 0.02 | 0.02 | 0.07 | 0.00 | 0.03 |
| **8** | 0.19 | 0.15 | 0.27 | 0.06 | 0.20 | 0.12 | 0.04 | 0.00 | 0.04 | 0.09 | 0.04 | 0.05 | 0.01 | 0.03 | 0.08 | 0.05 | 0.05 | 0.00 | 0.04 |
| **9** | 0.04 | 0.18 | 0.07 | 0.08 | 0.10 | 0.03 | 0.08 | 0.04 | 0.00 | 0.05 | 0.08 | 0.08 | -0.02 | 0.01 | 0.11 | -0.04 | 0.00 | 0.02 | 0.00 |
| **10** | 0.21 | 0.03 | 0.01 | 0.03 | 0.04 | 0.09 | 0.03 | 0.09 | 0.05 | 0.00 | 0.02 | 0.11 | 0.00 | 0.04 | 0.02 | 0.02 | 0.02 | 0.00 | 0.00 |
| **11** | -0.02 | 0.20 | 0.00 | 0.02 | 0.07 | 0.00 | 0.01 | 0.04 | 0.08 | 0.02 | 0.00 | 0.03 | 0.02 | 0.06 | -0.13 | -0.03 | 0.00 | 0.00 | -0.04 |
| **12** | -0.07 | 0.05 | 0.09 | 0.02 | 0.03 | 0.00 | 0.02 | 0.05 | 0.08 | 0.11 | 0.03 | 0.00 | 0.02 | 0.00 | -0.04 | 0.00 | 0.02 | 0.00 | 0.00 |
| **13** | 0.00 | 0.04 | 0.00 | 0.01 | 0.01 | 0.00 | 0.05 | 0.01 | -0.02 | 0.00 | 0.02 | 0.02 | 0.00 | 0.01 | 0.00 | 0.02 | 0.00 | 0.00 | 0.00 |
| **14** | 0.00 | 0.00 | 0.09 | 0.02 | 0.03 | 0.01 | 0.02 | 0.03 | 0.01 | 0.04 | 0.06 | 0.00 | 0.01 | 0.00 | 0.00 | 0.00 | 0.01 | -0.01 | 0.00 |
| **15** | 0.00 | 0.03 | 0.00 | 0.01 | 0.01 | 0.00 | 0.02 | 0.08 | 0.11 | 0.02 | -0.13 | -0.04 | 0.00 | 0.00 | 0.00 | 0.07 | 0.07 | -0.02 | -0.01 |
| **16** | -0.12 | 0.06 | 0.07 | 0.02 | 0.00 | 0.00 | 0.02 | 0.05 | -0.04 | 0.02 | -0.03 | 0.00 | 0.02 | 0.00 | 0.07 | 0.00 | -0.01 | 0.03 | 0.04 |
| **17** | 0.00 | 0.05 | 0.04 | 0.02 | 0.02 | 0.00 | 0.07 | 0.05 | 0.00 | 0.02 | 0.00 | 0.02 | 0.00 | 0.01 | 0.07 | -0.01 | 0.00 | 0.05 | 0.12 |
| **18** | 0.00 | 0.00 | 0.00 | 0.01 | 0.01 | 0.02 | 0.00 | 0.00 | 0.02 | 0.00 | 0.00 | 0.00 | 0.00 | -0.01 | -0.02 | 0.03 | 0.05 | 0.00 | 0.04 |
| **19** | 0.01 | 0.00 | 0.03 | 0.02 | 0.01 | 0.00 | 0.03 | 0.04 | 0.00 | 0.00 | -0.04 | 0.00 | 0.00 | 0.00 | -0.01 | 0.04 | 0.12 | 0.04 | 0.00 |
| **20** | 0.03 | 0.05 | 0.00 | 0.02 | 0.05 | 0.00 | 0.03 | 0.09 | -0.02 | -0.01 | -0.04 | 0.02 | 0.02 | 0.00 | 0.02 | 0.00 | 0.06 | 0.01 | 0.03 |
| **21** | 0.00 | 0.08 | 0.00 | 0.02 | 0.05 | 0.01 | 0.03 | 0.02 | 0.05 | 0.05 | 0.00 | 0.04 | 0.00 | 0.00 | 0.01 | -0.04 | 0.09 | 0.23 | 0.12 |
| **22** | -0.03 | 0.03 | 0.06 | 0.01 | 0.06 | 0.00 | 0.05 | 0.08 | 0.00 | 0.04 | -0.08 | 0.03 | -0.05 | -0.04 | -0.05 | -0.01 | 0.01 | 0.04 | 0.04 |
| **23** | 0.03 | 0.01 | 0.03 | 0.01 | 0.03 | 0.01 | 0.04 | 0.00 | 0.04 | 0.00 | 0.00 | 0.00 | 0.02 | -0.02 | 0.00 | -0.25 | 0.00 | 0.01 | 0.06 |
| **24** | -0.05 | 0.08 | 0.00 | 0.01 | 0.01 | 0.03 | 0.03 | 0.00 | 0.01 | 0.00 | -0.14 | -0.02 | -0.01 | 0.00 | -0.17 | 0.07 | 0.07 | 0.00 | 0.02 |
| **25** | 0.00 | 0.16 | 0.00 | 0.02 | 0.02 | 0.00 | 0.03 | 0.00 | 0.06 | 0.00 | -0.13 | -0.02 | 0.00 | 0.00 | -0.16 | 0.07 | 0.01 | 0.10 | 0.00 |
| **26** | 0.00 | 0.08 | 0.03 | 0.03 | 0.00 | 0.01 | 0.05 | 0.04 | -0.04 | 0.03 | 0.11 | 0.06 | 0.10 | -0.04 | 0.00 | -0.14 | -0.01 | 0.01 | -0.04 |

| | | | | | | | | | | | | | | | | | | | |
|---|---|---|---|---|---|---|---|---|---|---|---|---|---|---|---|---|---|---|---|
| **27** | -0.06 | 0.01 | 0.01 | 0.00 | 0.01 | 0.00 | 0.03 | 0.02 | 0.07 | 0.03 | 0.04 | 0.02 | 0.00 | 0.00 | -0.02 | -0.02 | 0.00 | -0.02 | 0.00 |
| **28** | 0.07 | 0.00 | 0.00 | 0.02 | 0.00 | 0.00 | 0.01 | 0.00 | 0.03 | -0.01 | 0.05 | 0.05 | 0.03 | 0.12 | 0.00 | 0.22 | -0.04 | 0.03 | 0.00 |
| **29** | 0.07 | 0.01 | 0.07 | 0.01 | 0.02 | 0.00 | 0.04 | 0.01 | -0.02 | 0.00 | -0.02 | -0.04 | 0.00 | -0.03 | -0.03 | 0.02 | 0.01 | 0.24 | 0.17 |
| **30** | 0.00 | 0.07 | 0.09 | 0.02 | 0.00 | 0.00 | 0.01 | 0.07 | 0.00 | 0.11 | -0.03 | 0.10 | 0.10 | 0.08 | -0.02 | 0.09 | -0.03 | 0.00 | -0.01 |
| **31** | -0.08 | 0.11 | 0.02 | 0.01 | 0.02 | 0.00 | 0.07 | 0.08 | -0.06 | -0.11 | 0.07 | 0.03 | 0.00 | 0.04 | 0.00 | 0.01 | 0.00 | 0.01 | -0.01 |
| **32** | 0.01 | 0.00 | 0.07 | 0.04 | 0.00 | 0.01 | 0.03 | 0.03 | -0.04 | -0.02 | 0.00 | 0.03 | 0.02 | -0.04 | 0.00 | 0.00 | 0.00 | -0.03 | 0.00 |
| **33** | -0.03 | 0.03 | 0.01 | 0.02 | 0.01 | 0.04 | 0.06 | 0.03 | 0.03 | 0.00 | 0.02 | 0.07 | 0.02 | -0.03 | -0.10 | 0.16 | 0.05 | 0.19 | -0.01 |
| **34** | 0.07 | 0.03 | 0.07 | 0.01 | 0.10 | 0.05 | 0.01 | 0.03 | -0.06 | 0.00 | 0.01 | 0.02 | 0.03 | 0.01 | 0.03 | 0.00 | 0.04 | -0.01 | 0.03 |
| **35** | 0.01 | 0.08 | 0.01 | 0.02 | 0.02 | 0.00 | 0.01 | 0.02 | -0.02 | 0.00 | 0.04 | 0.02 | 0.04 | 0.26 | 0.06 | 0.21 | 0.00 | 0.15 | 0.04 |
| **36** | -0.08 | 0.06 | 0.02 | 0.05 | 0.02 | 0.07 | 0.08 | 0.03 | -0.03 | -0.10 | 0.01 | 0.00 | -0.02 | 0.02 | 0.01 | 0.06 | 0.02 | 0.01 | 0.02 |
| **37** | 0.12 | 0.03 | 0.09 | 0.01 | 0.03 | 0.04 | 0.01 | 0.11 | -0.02 | 0.05 | 0.08 | 0.00 | 0.15 | -0.02 | 0.01 | -0.06 | 0.00 | 0.00 | 0.00 |
| **38** | -0.01 | 0.04 | 0.02 | 0.04 | 0.01 | 0.06 | 0.04 | 0.01 | -0.02 | -0.01 | 0.07 | 0.04 | 0.03 | 0.03 | 0.05 | -0.02 | 0.13 | 0.05 | -0.03 |

| | **20** | **21** | **22** | **23** | **24** | **25** | **26** | **27** | **28** | **29** | **30** | **31** | **32** | **33** | **34** | **35** | **36** | **37** | **38** |
|---|---|---|---|---|---|---|---|---|---|---|---|---|---|---|---|---|---|---|---|
| **1** | 0.03 | 0.00 | -0.03 | 0.03 | -0.05 | 0.00 | 0.00 | -0.06 | 0.07 | 0.07 | 0.00 | -0.08 | 0.01 | -0.03 | 0.07 | 0.01 | -0.08 | 0.12 | -0.01 |
| **2** | 0.05 | 0.08 | 0.03 | 0.01 | 0.08 | 0.16 | 0.08 | 0.01 | 0.00 | 0.01 | 0.07 | 0.11 | 0.00 | 0.03 | 0.03 | 0.08 | 0.06 | 0.03 | 0.04 |
| **3** | 0.00 | 0.00 | 0.06 | 0.03 | 0.00 | 0.00 | 0.03 | 0.01 | 0.00 | 0.07 | 0.09 | 0.02 | 0.07 | 0.01 | 0.07 | 0.01 | 0.02 | 0.09 | 0.02 |
| **4** | 0.02 | 0.02 | 0.01 | 0.01 | 0.01 | 0.02 | 0.03 | 0.00 | 0.02 | 0.01 | 0.02 | 0.01 | 0.04 | 0.02 | 0.01 | 0.02 | 0.05 | 0.01 | 0.04 |
| **5** | 0.05 | 0.05 | 0.06 | 0.03 | 0.01 | 0.02 | 0.00 | 0.01 | 0.00 | 0.02 | 0.00 | 0.02 | 0.00 | 0.01 | 0.10 | 0.02 | 0.02 | 0.03 | 0.01 |
| **6** | 0.00 | 0.01 | 0.00 | 0.01 | 0.03 | 0.00 | 0.01 | 0.00 | 0.00 | 0.00 | 0.00 | 0.00 | 0.01 | 0.04 | 0.05 | 0.00 | 0.07 | 0.04 | 0.06 |
| **7** | 0.03 | 0.03 | 0.05 | 0.04 | 0.03 | 0.03 | 0.05 | 0.03 | 0.01 | 0.04 | 0.01 | 0.07 | 0.03 | 0.06 | 0.01 | 0.01 | 0.08 | 0.01 | 0.04 |
| **8** | 0.09 | 0.02 | 0.08 | 0.00 | 0.00 | 0.00 | 0.04 | 0.02 | 0.00 | 0.01 | 0.07 | 0.08 | 0.03 | 0.03 | 0.03 | 0.02 | 0.03 | 0.11 | 0.01 |
| **9** | -0.02 | 0.05 | 0.00 | 0.04 | 0.01 | 0.06 | -0.04 | 0.07 | 0.03 | -0.02 | 0.00 | -0.06 | -0.04 | 0.03 | -0.06 | -0.02 | -0.03 | -0.02 | -0.02 |
| **10** | -0.01 | 0.05 | 0.04 | 0.00 | 0.00 | 0.00 | 0.03 | 0.03 | -0.01 | 0.00 | 0.11 | -0.11 | -0.02 | 0.00 | 0.00 | 0.00 | -0.10 | 0.05 | -0.01 |
| **11** | -0.04 | 0.00 | -0.08 | 0.00 | -0.14 | -0.13 | 0.11 | 0.04 | 0.05 | -0.02 | -0.03 | 0.07 | 0.00 | 0.02 | 0.01 | 0.04 | 0.01 | 0.08 | 0.07 |
| **12** | 0.02 | 0.04 | 0.03 | 0.00 | -0.02 | -0.02 | 0.06 | 0.02 | 0.05 | -0.04 | 0.10 | 0.03 | 0.03 | 0.07 | 0.02 | 0.02 | 0.00 | 0.00 | 0.04 |

| | | | | | | | | | | | | | | | | | | | |
|---|---|---|---|---|---|---|---|---|---|---|---|---|---|---|---|---|---|---|---|
| **13** | 0.02 | 0.00 | -0.05 | 0.02 | -0.01 | 0.00 | 0.10 | 0.00 | 0.03 | 0.00 | 0.10 | 0.00 | 0.02 | 0.02 | 0.03 | 0.04 | -0.02 | 0.15 | 0.03 |
| **14** | 0.00 | 0.00 | -0.04 | -0.02 | 0.00 | 0.00 | -0.04 | 0.00 | 0.12 | -0.03 | 0.08 | 0.04 | -0.04 | -0.03 | 0.01 | 0.26 | 0.02 | -0.02 | 0.03 |
| **15** | 0.02 | 0.01 | -0.05 | 0.00 | -0.17 | -0.16 | 0.00 | -0.02 | 0.00 | -0.03 | -0.02 | 0.00 | 0.00 | -0.10 | 0.03 | 0.06 | 0.01 | 0.01 | 0.05 |
| **16** | 0.00 | -0.04 | -0.01 | -0.25 | 0.07 | 0.07 | -0.14 | -0.02 | 0.22 | 0.02 | 0.09 | 0.01 | 0.00 | 0.16 | 0.00 | 0.21 | 0.06 | -0.06 | -0.02 |
| **17** | 0.06 | 0.09 | 0.01 | 0.00 | 0.07 | 0.01 | -0.01 | 0.00 | -0.04 | 0.01 | -0.03 | 0.00 | 0.00 | 0.05 | 0.04 | 0.00 | 0.02 | 0.00 | 0.13 |
| **18** | 0.01 | 0.23 | 0.04 | 0.01 | 0.00 | 0.10 | 0.01 | -0.02 | 0.03 | 0.24 | 0.00 | 0.01 | -0.03 | 0.19 | -0.01 | 0.15 | 0.01 | 0.00 | 0.05 |
| **19** | 0.03 | 0.12 | 0.04 | 0.06 | 0.02 | 0.00 | -0.04 | 0.00 | 0.00 | 0.17 | -0.01 | -0.01 | 0.00 | -0.01 | 0.03 | 0.04 | 0.02 | 0.00 | -0.03 |
| **20** | 0.00 | 0.06 | 0.04 | 0.00 | 0.00 | -0.01 | 0.00 | -0.04 | 0.00 | -0.03 | 0.05 | -0.03 | 0.00 | 0.02 | 0.13 | 0.00 | 0.08 | 0.07 | 0.00 |
| **21** | 0.06 | 0.00 | 0.05 | -0.02 | 0.07 | 0.05 | 0.00 | -0.02 | -0.01 | 0.00 | -0.02 | 0.01 | 0.02 | 0.05 | 0.02 | 0.03 | 0.07 | 0.03 | 0.04 |
| **22** | 0.04 | 0.05 | 0.00 | -0.03 | -0.07 | -0.06 | 0.01 | 0.00 | -0.04 | -0.03 | 0.02 | 0.00 | 0.03 | 0.00 | 0.06 | 0.02 | 0.06 | 0.08 | 0.02 |
| **23** | 0.00 | -0.02 | -0.03 | 0.00 | 0.03 | 0.16 | 0.00 | 0.00 | 0.16 | -0.03 | 0.00 | 0.03 | 0.00 | -0.07 | -0.01 | 0.24 | 0.00 | -0.02 | 0.03 |
| **24** | 0.00 | 0.07 | -0.07 | 0.03 | 0.00 | -0.29 | 0.00 | 0.00 | 0.13 | 0.02 | -0.07 | 0.01 | 0.00 | -0.02 | 0.00 | 0.02 | 0.02 | 0.00 | 0.08 |
| **25** | -0.01 | 0.05 | -0.06 | 0.16 | -0.29 | 0.00 | 0.05 | 0.00 | -0.03 | -0.02 | 0.00 | 0.02 | 0.00 | 0.00 | -0.02 | 0.00 | 0.01 | -0.03 | 0.10 |
| **26** | 0.00 | 0.00 | 0.01 | 0.00 | 0.00 | 0.05 | 0.00 | 0.00 | 0.06 | 0.00 | 0.03 | 0.05 | 0.02 | 0.05 | 0.00 | 0.06 | 0.02 | -0.29 | 0.04 |
| **27** | -0.04 | -0.02 | 0.00 | 0.00 | 0.00 | 0.00 | 0.00 | 0.00 | 0.02 | 0.00 | 0.00 | 0.06 | 0.03 | 0.00 | 0.04 | 0.00 | 0.00 | 0.00 | 0.09 |
| **28** | 0.00 | -0.01 | -0.04 | 0.16 | 0.13 | -0.03 | 0.06 | 0.02 | 0.00 | 0.00 | 0.19 | 0.03 | 0.02 | -0.02 | 0.01 | -0.11 | 0.01 | 0.00 | 0.03 |
| **29** | -0.03 | 0.00 | -0.03 | -0.03 | 0.02 | -0.02 | 0.00 | 0.00 | 0.00 | 0.00 | 0.00 | 0.00 | 0.00 | -0.07 | 0.04 | 0.05 | -0.02 | 0.02 | -0.06 |
| **30** | 0.05 | -0.02 | 0.02 | 0.00 | -0.07 | 0.00 | 0.03 | 0.00 | 0.19 | 0.00 | 0.00 | 0.03 | 0.06 | -0.02 | 0.00 | 0.09 | 0.00 | 0.00 | 0.00 |
| **31** | -0.03 | 0.01 | 0.00 | 0.03 | 0.01 | 0.02 | 0.05 | 0.06 | 0.03 | 0.00 | 0.03 | 0.00 | 0.06 | 0.00 | -0.03 | 0.02 | -0.13 | 0.00 | -0.04 |

| | | | | | | | | | | | | | | | | | | | |
|---|---|---|---|---|---|---|---|---|---|---|---|---|---|---|---|---|---|---|---|
| **32** | 0.00 | 0.02 | 0.03 | 0.00 | 0.00 | 0.00 | 0.02 | 0.03 | 0.02 | 0.00 | 0.06 | 0.06 | 0.00 | 0.00 | 0.00 | 0.02 | 0.02 | 0.02 | 0.04 |
| **33** | 0.02 | 0.05 | 0.00 | -0.07 | -0.02 | 0.00 | 0.05 | 0.00 | -0.02 | -0.07 | -0.02 | 0.00 | 0.00 | 0.00 | 0.02 | -0.05 | 0.03 | 0.02 | 0.00 |
| **34** | 0.13 | 0.02 | 0.06 | -0.01 | 0.00 | -0.02 | 0.00 | 0.04 | 0.01 | 0.04 | 0.00 | -0.03 | 0.00 | 0.02 | 0.00 | 0.00 | 0.03 | 0.03 | -0.04 |
| **35** | 0.00 | 0.03 | 0.02 | 0.24 | 0.02 | 0.00 | 0.06 | 0.00 | -0.11 | 0.05 | 0.09 | 0.02 | 0.02 | -0.05 | 0.00 | 0.00 | 0.00 | -0.01 | 0.03 |
| **36** | 0.08 | 0.07 | 0.06 | 0.00 | 0.02 | 0.01 | 0.02 | 0.00 | 0.01 | -0.02 | 0.00 | -0.13 | 0.02 | 0.03 | 0.03 | 0.00 | 0.00 | 0.04 | -0.01 |
| **37** | 0.07 | 0.03 | 0.08 | -0.02 | 0.00 | -0.03 | -0.29 | 0.00 | 0.00 | 0.02 | 0.00 | 0.00 | 0.02 | 0.02 | 0.03 | -0.01 | 0.04 | 0.00 | 0.02 |
| **38** | 0.00 | 0.04 | 0.02 | 0.03 | 0.08 | 0.10 | 0.04 | 0.09 | 0.03 | -0.06 | 0.00 | -0.04 | 0.04 | 0.00 | -0.04 | 0.03 | -0.01 | 0.02 | 0.00 |

1-Age, 2-Sex, 3-Household food security status, 4-Income, 5-Education, 6-Worked in the past week, 7-Geography, 8-Smoking status, 9-Alcohol, 10-Coffee/tea, 11-Cured meat, 12-Dairy (higher fat), 13-Dairy (lower fat), 14-Eggs, 15-Fish, 16-Grains (other), 17-Green vegetables, 18-Onion/garlic, 19-Orange vegetables, 20-Other fruits, 21-Other vegetables, 22-Plant proteins, 23-Potato, 24-Poultry, 25-Red meat, 26-Refined grains, 27-Salty snacks, 28-Saturated fats, 29-Soup, 30-Sugar, 31-Sweet beverages, 32-Sweet treats, 33-Tomato, 34-Tropical fruits, 35-Unsaturated fats, 36-Water, 37-Whole grains, 38-Other food

### Reporting checklist for observational studies in nutritional epidemiology.

Based on the STROBE-nut guidelines.

#### Instructions to authors

Complete this checklist by entering the page numbers from your manuscript where readers will find each of the items listed below.

Your article may not currently address all the items on the checklist. Please modify your text to include the missing information. If you are certain that an item does not apply, please write "n/a" and provide a short explanation.

Upload your completed checklist as an extra file when you submit to a journal.

In your methods section, say that you used the STROBE-nutreporting guidelines, and cite them as:

Lachat C, Hawwash D, Ocké MC, Berg C, Forsum E, Hörnell A, Larsson C, Sonestedt E, Wirfält E, Åkesson A, Kolsteren P, Byrnes G, De Keyzer W, Van Camp J, Cade JE, Slimani N, Cevallos M, Egger M, Huybrechts I. Strengthening the Reporting of Observational Studies in Epidemiology-Nutritional Epidemiology (STROBE-nut): An Extension of the STROBE Statement.

| | | Reporting Item | Page Number |
|---|---|---|---|
| **Title and abstract** | | | |
| Title | #1a | Indicate the study’s design with a commonly used term in the title or the abstract | 1 |
| None | #nut-1 | State the dietary/nutritional assessment method(s) used in the title or in the abstract. | 3 |
| Abstract | #1b | Provide in the abstract an informative and balanced summary of what was done and what was found | 3 |
| **Introduction** | | | |
| Background / rationale | #2 | Explain the scientific background and rationale for the investigation being reported | 5-8 |
| Objectives | #3 | State specific objectives, including any prespecified hypotheses | 8 |
| **Methods** | | | |
| Study design | #4 | Present key elements of study design early in the paper | 8 |
| Setting | #5 | Describe the setting, locations, and relevant dates, including periods of recruitment, exposure, follow-up, and data collection | 8-9 |
| None | #nut-5 | Describe any characteristics of the study settings that might affect the dietary intake or nutritional status of the participants, if applicable. | 8-9 |
| Eligibility | #6a | Cohort study: Give the eligibility criteria and the sources and methods of selection of participants. Describe methods of follow-up. Case-control study: Give the eligibility criteria and the sources and methods of case ascertainment and control selection. Give the rationale for the choice of cases and controls. Cross-sectional study: Give the eligibility criteria, and the sources and methods of selection of participants. | 8-9 |

| | | | |
|---|---|---|---|
| None | #nut-6 | Report any particular dietary, physiologic, or nutritional characteristics that were considered when selecting the target population. | 9 |
| None | #6b | Cohort study: For matched studies, give matching criteria and number of exposed and unexposed. Case-control study: For matched studies, give matching criteria and the number of controls per case. | N/A |
| Variables | #7 | Clearly define all outcomes, exposures, predictors, potential confounders, and effect modifiers. Give diagnostic criteria, if applicable | 9-10 |
| None | #nut-7.1 | Clearly define foods, food groups, nutrients, or other food components (e.g., preparation method, taxonomical descriptors, classification, chemical form). | 10-11 |
| None | #nut-7.2 | When calculating dietary patterns, describe the methods to obtain them and their nutritional properties. | 11-13 |
| Data sources and measurement | #8 | For each variable of interest give sources of data and details of methods of assessment (measurement). Describe comparability of assessment methods if there is more than one group. Give information separately for for exposed and unexposed groups if applicable. | 10-11 |
| None | #nut-8.1 | Describe the dietary assessment method(s) (e.g., portion size estimation, number of days and items recorded, how it was developed and administered, and how quality was ensured); report if and how supplement intake was assessed. | 10-11 |
| None | #nut-8.2 | Describe and justify food-composition data used; explain the procedure to match food composition with consumption data; describe the use of conversion factors, if applicable | 11 |
| None | #nut-8.3 | Describe the nutrient requirements, recommendations, or dietary guidelines and the evaluation approach used to compare intake with the dietary reference values, if applicable | N/A |
| None | #nut-8.4 | When using nutritional biomarkers, additionally use the STROBE-ME; report the type of biomarkers used and usefulness as dietary exposure markers | N/A |
| None | #nut-8.5 | Describe the assessment of nondietary data (e.g., nutritional status and influencing factors) and timing of the assessment of these variables in relation to dietary assessment | 8 |
| None | #nut-8.6 | Report on the validity of the dietary or nutritional assessment methods and any internal or external validation used in the study, if applicable | N/A |
| Bias | #9 | Describe any efforts to address potential sources of bias | 9, 22-23 |
| None | #nut-9 | Report how bias in dietary or nutritional assessment was addressed (e.g., misreporting, changes in habits as a result of being measured, data imputation from other sources). | 9-11 |
| Study size | #10 | Explain how the study size was arrived at | 9 |
| Quantitative variables | #11 | Explain how quantitative variables were handled in the analyses. If applicable, describe which groupings were chosen, and why | 9-10 |

| | | | |
|---|---|---|---|
| None | #nut-11 | Explain categorization of dietary/nutritional data (e.g., use of N-tiles and handling of nonconsumers) and the choice of reference category, if applicable. | 10-11 |
| Statistical methods | #12a | Describe all statistical methods, including those used to control for confounding | 11-13 |
| Subgroups and interactions | #12b | Describe any methods used to examine subgroups and interactions | 12-13 |
| Missing data | #12c | Explain how missing data were addressed | 9 |
| Loss to follow up | #12d | Cohort study: if applicable, explain how loss to follow-up was addressed. Case-control study: if applicable, explain how matching of cases and controls was addressed. Cross-sectional study: if applicable, describe analytical methods taking account of sampling strategy. | 8-9 |
| Sensitivity analysis | #12e | Describe any sensitivity analyses | 9, 22 |
| None | #nut-12.1 | Describe any statistical method used to combine dietary or nutritional data, if applicable. | N/A |
| None | #nut-12.2 | Describe and justify the method for energy adjustments, intake modeling, and use of weighting factors, if applicable | 11, 13 |
| None | #nut-12.3 | Report any adjustments for measurement error (i.e., from a validity or calibration study). | N/A |
| **Results** | | | |
| Participants | #13a | Report numbers of individuals at each stage of study—eg numbers potentially eligible, examined for eligibility, confirmed eligible, included in the study, completing follow-up, and analysed. Give information separately for for exposed and unexposed groups if applicable. | 8-9 |
| Non-participation | #13b | Give reasons for non-participation at each stage | N/A |
| Participant journey | #13c | Consider the use of a flow diagram | N/A |
| None | #nut-13 | Report the number of individuals excluded on the basis of missing, incomplete, or implausible dietary and nutritional data. | 9 |
| Descriptive data | #14a | Give characteristics of study participants (eg demographic, clinical, social) and information on exposures and potential confounders. Give information separately for exposed and unexposed groups if applicable. | 13, 33 |
| Missing data | #14b | Indicate number of participants with missing data for each variable of interest | 9 |
| Follow-up time | #14c | Cohort study: Summarise follow-up time (eg, average and total amount) | N/A |

| | | | |
|---|---|---|---|
| None | #nut-14 | Give the distribution of participant characteristics across the exposure variables, if applicable; specify if food consumption for the total population or consumers only was used to obtain results | N/A |
| Outcome data | #15 | Cohort study: report numbers of outcome events or summary measures over time. Case-control study: report numbers in each exposure category, or summary measures of exposure. Cross-sectional study: report numbers of outcome events or summary measures. | N/A |
| Main results | #16a | Give unadjusted estimates and, if applicable, confounder-adjusted estimates and their precision (eg, 95% confidence interval). Make clear which confounders were adjusted for and why they were included | 14-17, 34-38 |
| Category boundaries | #16b | Report category boundaries when continuous variables were categorized | N/A |
| Relative and absolute risks | #16c | If relevant, consider translating estimates of relative risk into absolute risk for a meaningful time period | N/A |
| None | #nut-16 | Specify if nutrient intakes are reported with or without the inclusion of dietary supplement intake, if applicable. | N/A |
| Other analyses | #17 | Report other analyses done—eg analyses of subgroups and interactions, and sensitivity analyses | 14-17 |
| None | #nut-17 | Report any sensitivity analysis (e.g., exclusion of misreporters or outliers) and data imputation, if applicable | N/A |
| **Discussion** | | | |
| Key results | #18 | Summarise key results with reference to study objectives | 17 |
| Limitations | #19 | Discuss limitations of the study, taking into account sources of potential bias or imprecision. Discuss both direction and magnitude of any potential bias. | 22-23 |
| None | #nut-19 | Describe the main limitations of the data sources and assessment methods used and implications for the interpretation of the findings | 22-23 |
| Interpretation | #20 | Give a cautious overall interpretation considering objectives, limitations, multiplicity of analyses, results from similar studies, and other relevant evidence. | 18-24 |
| None | #nut-20 | Report the nutritional relevance of the findings, given the complexity of diet or nutrition as an exposure. | 18-21 |
| Generalisability | #21 | Discuss the generalisability (external validity) of the study results | 18-19 |
| **Other Information** | | | |
| Funding | #22 | Give the source of funding and the role of the funders for the present study and, if applicable, for the original study on which the present article is based | 1 |
| Ethics | #nut-22.1 | Describe the procedure for consent and study approval from ethics committee(s). | 9 |
| Data statement | #nut-22.2 | Provide data collection tools and data as online material or explain how they can be accessed | 25 |

None The STROBE-nut checklist is distributed under the terms of the Creative Commons Attribution License CC-BY. This checklist can be completed online using https://www.goodreports.org/, a tool made by the EQUATOR Network in collaboration with Penelope.ai
